# Tanycytes are degraded in Alzheimer’s Disease, disrupting the brain-to-blood efflux of Tau

**DOI:** 10.1101/2022.05.04.22274181

**Authors:** Florent Sauvé, Gaëtan Ternier, Julie Dewisme, Thibaut Lebouvier, Elian Dupré, Clément Danis, S. Rasika, Young-Bum Kim, Philippe Ciofi, Paolo Giacobini, Luc Buée, Isabelle Landrieu, Florence Pasquier, Claude-Alain Maurage, Ruben Nogueiras, Markus Schwaninger, Vincent Prevot

**Author notes:** Data used in preparation of this article were obtained from the Alzheimer’s Disease Neuroimaging Initiative (ADNI) database (adni.loni.usc.edu). As such, the investigators within the ADNI contributed to the design and implementation of ADNI and/or provided data but did not participate in analysis or writing of this report. A complete listing of ADNI investigators can be found at: http://adni.loni.usc.edu/wpcontent/uploads/how_to_apply/ADNI_Acknowledgement_List.pdf. These authors jointly supervised this work (Ruben Nogueiras, Markus Schwaninger, Vincent Prevot). Corresponding author:* Vincent Prevot, Ph.D., Inserm U1172, Bâtiment Biserte, Place de Verdun, 59045 Lille Cedex, France.

## Abstract

The accumulation of pathological Tau in the brain and cerebrospinal fluid (CSF) and its eventual increase in the blood are hallmarks of Alzheimer’s disease (AD). However, the mechanisms of Tau clearance from the brain to the periphery are not clear. We show here, using animal and cellular models as well as patient blood samples and post mortem brains, that hypothalamic tanycytes, whose cell bodies line the ventricular wall and send long processes to the underlying pituitary portal capillary bed, take up and transport Tau from the CSF and release it into these capillaries, whence it travels to the pituitary and eventually the systemic circulation. Specifically blocking tanycytic vesicular transport leads to an accumulation of exogenous fluorescent Tau in the CSF of mice. In AD and frontotemporal dementia, tanycytic morphology is altered, with a dramatic fragmentation of the secondary cytoskeleton in the former but not the latter, accounting for reduced CSF Tau clearance in AD. Both the implication of tanycytic degradation in the pathophysiology of a human disease and the evidence for the existence of a brain-to-blood tanycytic shuttle are unprecedented, and raise important questions regarding the role of tanycytes in physiological clearance mechanisms and the development of neurodegenerative disorders.

## INTRODUCTION

In our ageing societies, Alzheimer’s Disease (AD) and other age-related dementias such as frontotemporal dementia (FTD) have become a major public health issue (*1*). The pathogenesis of AD is characterized by the accumulation and aggregation of amyloid beta peptide (Aβ) and the microtubule-associated protein Tau in the brain, which trigger neuronal dysfunction and death (*2–8*). Although intracellular, these proteins are released from cells and subsequently cleared from the brain under physiological conditions (*9, 10*). The impairment of the mechanisms responsible for clearing these pathogenic proteins from the brain, either by degradation or by transport out of the brain, contribute to the neuronal pathology (*11*). The cerebrospinal fluid (CSF) is thought to be an intermediary destination in the clearance of Tau from the brain parenchyma or interstitial fluid (*12, 13*), and Tau protein concentrations are increased in the CSF of AD patients (*14–16*). However, while CSF-borne Tau is seen to travel to the blood (*17, 18*) and increased circulating Tau levels are even considered a putative biomarker for AD (*19*), the cellular mechanisms underlying this transfer of Tau from the CSF to the blood remain unknown.

The brain-to-blood transfer of molecules is regulated by two distinct barriers, the traditional blood-brain barrier (BBB), consisting of the endothelium of the brain vasculature, and the blood-CSF barrier (BCSFB), located at the choroid plexus and the circumventricular organs (CVO) (*20*). CVOs are peculiar brain structures characterized by the presence of a highly permeable vasculature due to the fenestration of the endothelium (*21–23*). One of these organs, the median eminence (ME) of the hypothalamus, has been extensively documented as a hotspot for brain/blood exchanges in the context of metabolism and the hormonal axis (*24, 25*). In the ME, the barrier function is displaced from endothelial cells to specialized ependymoglial cells called tanycytes (*26*), characterized by long slim processes stretching from the wall of the cerebral ventricle, where their cell bodies are located, to the fenestrated capillaries of the ME, which connect the hypothalamus to the anterior pituitary and thereby to the general circulation. Tanycytes, which constitute the BCSFB in the ME and serve as a bridge between the CSF and the pituitary portal circulation, perform several key functions (reviewed in (*27*)), including the active shuttling of blood-borne signals into the CSF (*28–32*). Whether tanycytes are also endowed with the ability to shuttle molecules in the reverse direction, from the CSF to ME capillaries, and whether such a mechanism could be of pathophysiological relevance in AD or other dementias, have not been formally explored. Here, we investigated the ability of tanycytes to take up and release Tau in a preclinical setting both *in vitro* and *in vivo*, and examined the implications of this tanycytic transport for Tau clearance and AD pathology using post mortem human brains and an AD patient database.

## RESULTS

### Tanycytes are Tau transporters

In order to determine whether Tau could be transported by tanycytes, we first examined its uptake by rat tanycytes in primary culture. To be able to track Tau internalization, we conjugated 2N4R Tau to a fluorescent tag (Tau-565), resulting in a mix of Tau molecules carrying 1 to 5 fluorophore tags (Supplementary Fig. 1A). The incubation of primary rat tanycytes with Tau-565 for 30 minutes led to the visualization of intracellular Tau-positive vesicles, as shown by both Tau-565 fluorescence and immunolabeling using the Tau5 antibody (Fig. 1A). Western blotting of untreated rat tanycytes revealed no detectable Tau protein, but a 30-minute treatment with an unconjugated human recombinant 2N4R Tau isoform led to a positive signal on the blots (Supplementary Fig. 1B), suggesting that Tau conjugation has no effect on its uptake. Moreover, the presence of clathrin on Tau-565-positive vesicles suggest the involvement of clathrin-coated vesicles in Tau endocytosis (Supplementary Fig. 1C) Interestingly, live imaging revealed that these intracellular Tau-565-positive vesicles were in motion (Video 1). To verify whether this intracellular movement meant that internalized Tau was being exocytosed by tanycytes, we incubated rat primary tanycytes with Tau-565 for 30 minutes, and used an ELISA specific to human Tau to measure its levels in fresh medium at various time points. Primary tanycytes showed a rapid release of Tau that increased over time, supporting the view that Tau uptake by tanycytes is the first step in its tanycytic clearance (Fig 1B).

**Figure 1:**
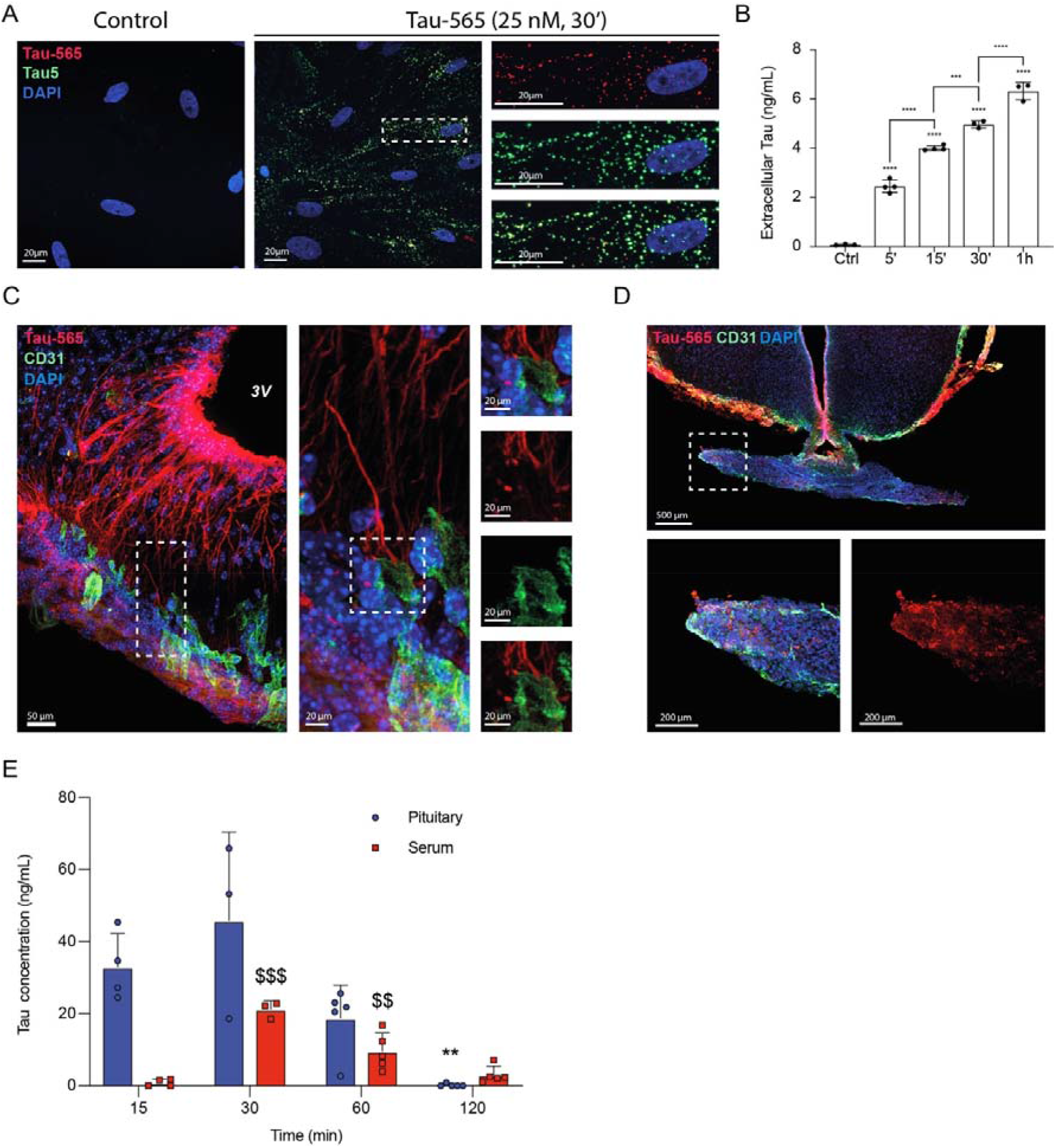
Tanycytes take up and secrete Tau *in vitro* and *in vivo*. A. Photomicrograph of cultured rat primary tanycytes treated or not with Tau-565 (red) and immunolabeled for Tau (Tau5 antibody, green). B. ELISA measurement of secreted human Tau in tanycyte primary culture medium at different time points following incubation with and uptake of Tau-565 (n = 3, 4, 4, 3 and 3 wells of one primary culture). One-way analysis of variance (ANOVA) (p<0.0001, F_(4, 12)_= 406) with Tukey’s multiple comparison post-hoc test (Ctrl vs. 5’: q_(12)_=21.11, p<0.0001; Ctrl vs. 15’: q_(12)_=34.8; Ctrl vs 30’: q_(12)_=40.58, p<0.0001; Ctrl vs. 1h: q_(12)_=51,78, p<0.0001; 5’ vs. 15’: q_(12)_=14.59, p<0.0001; 15’ vs. 30’: q_(12)_=8.573, p=0.004; 30’ vs. 60’: q_(12)_=11.2, p<0.001). Data are presented as means ± S.D., ***, p<0.001, ****, p<0.0001. C. Representative photomicrograph of the median eminence of aTau-565 (red)-injected wild-type (WT) mouse immunolabeled for CD31 (green) to visualize blood vessels. D. Photomicrograph of the median eminence and attached pituitary of aTau-565 (red)-injected WT mouse immunolabeled for CD31 (green) to visualize blood vessels. E. ELISA measurement of human Tau in the pituitary and serum of Tau-565-injected WT mice (n=4, 3, 5 and 5 mice per time point). Data were analyzed separately for the pituitary and serum using one-way ANOVA (pituitary: F_(3,13)_=11.13, p=0.0007; serum: F_(3,13)_=29.86, p<0.001) followed by Dunn’s multiple comparison post-hoc test (pituitary: 15’ vs. 2h, q_(13)_=5.873, p=0.0055; serum: 15’ vs. 30’, q_(13)_=11.38, p<0.001; 15’ vs. 1h, q_(13)_=5.531, p=0.008). Data are presented as means ± S.D. **, p<0.01 for the pituitary; $$, p<0.01 and $$$, p<0.001 for the serum.

Since tanycytes appear to both take up and release Tau *in vitro*, we next investigated the time course and destination of this Tau cleared from the CSF *in vivo*, using wild-type mice. We performed ICV injections of Tau-565 into the lateral ventricle, followed by immunohistofluorescence and ELISA measurement of Tau in the pituitary, to which the portal capillaries of the ME lead, and in the general circulation. In the brain, Tau was observed at the luminal surface of the ventricular wall at all time points (Supplementary Fig. 1D). However, while this labeling remained faint and restricted to the lumen in other areas bordering the ventricles, tanycytes of the ME, as revealed by vimentin immunolabeling, were strongly positive for Tau-565 from their cell bodies at the ventricular wall to their endfeet in contact with the fenestrated vessels of the ME, as early as 15 minutes post-injection (Fig. 1C), corroborating our observations in rat tanycytes *in vitro*. This Tau accumulation by tanycytic cell bodies and its intracellular transport could no longer be seen by 1 hour post-injection (Supplementary Fig. 1D), reminiscent of its exocytosis over time by primary cultured tanycytes above. The distribution of Tau-565 along the third ventricle as a whole was also examined using tissue clearing and light-sheet imaging of Tau-565 injected brains 30 minutes after ICV Tau-565 injection. The Tau-565 signal was found to be concentrated along the ME, especially in ventral tanycytes (Video 2, Supplementary Fig. 1E), further supporting a key role for these cells in trapping and clearing Tau from the brain. To verify the specificity of Tau uptake and secretion by tanycytes compared to other proteins, we performed ICV injections of fluorophore-coupled bovine serum albumin (BSA-565), which has a molecular weight of the same order of magnitude as Tau and bovine or human serum albumin have been used as a control for Tau in previous studies (*33, 34*); 30 minutes post-injection, the BSA-565 signal was detected along the lumen of the third ventricle, similar to Tau-565 in the rest of the brain, but not in tanycytic cell bodies or endfeet (Supplementary Fig. 1F).

Next, we studied the route and kinetics of Tau-565 transport from the brain to the systemic blood circulation. The ME vasculature being part of the pituitary portal circulation, we could expect Tau exocytosed by tanycytes to follow a pituitary route. Indeed, mouse pituitaries were positive for Tau-565 fluorescence 30 minutes after injection, further supporting a role for ME tanycytes as major Tau transporters, and the sequential release of Tau by tanycytic endfeet into the portal circulation and its transport to the pituitary (Fig. 1D). In accordance with this observation, recombinant human Tau was detected in pituitary tissue by ELISA starting 15 minutes post-injection and up to 1h, while it was detectable in peripheral blood serum by 30 minutes post-injection, decreasing over time until the second hour post-injection (Fig. 1E). This temporality of Tau detection in the brain, pituitary and blood also supports its transport through ME tanycytes to the portal circulation for delivery to the pituitary, and eventually to the systemic circulation.

### Impairing tanycytic transport blunts brain-to-blood Tau efflux

In order to investigate the specific role of tanycytes in Tau efflux from the brain to the blood, we used a murine model in which we arrested vesicular transport in tanycytes using the botulinum toxin light chain B (BoNT/B). BoNT/B prevents exocytosis in neurons and glial cells by cleaving vesicle-associated membrane proteins 1 and 2 (VAMPs), which mediate vesicular fusion and transport (*35, 36*). Another BoNT/B target, VAMP3, is also implicated in endocytosis in platelets (*37*). BoNT/B expression in tanycytes could thus be used to impair the uptake, transport and release of various substances transported by tanycytes, including Tau. Transgenic mice expressing a floxed BoNT/B, or iBot mice (*35*), were infected with an AAV1/2-Dio2-iCre-GFP virus to induce Cre expression specifically in tanycytes (Fig. 2A, B). The infection rate of tanycytes was 71% for ventral tanycytes and 41% for dorsal tanycytes (Fig. 2C). After ICV Tau-565 injection, iBot mice showed a dramatic reduction in the Tau-565 signal in tanycytic processes and endfeet as well as the underlying capillary bed compared to controls (Fig. 2D), linked to reduced Tau-565 uptake by many infected tanycytic cell bodies at the ventricular wall (Fig. 2E). Correspondingly, 30 minutes post-injection, pituitary concentrations of Tau were almost halved in iBot mice (n=5, 26.65 ± 8.22 ng/mL) compared to control mice (n=4, 47.53 ± 11.97 ng/mL, p=0.004, Fig. 2F), whereas plasma Tau concentrations underwent a 3.6-fold reduction, from 16.44 ± 6.66 ng/mL to 4.55 ± 2.85 ng/mL (n=8 and n=10, p<0.001, Fig. 2G), emphasizing the importance of tanycytic transport in Tau efflux from the brain to the blood.

**Figure 2:**
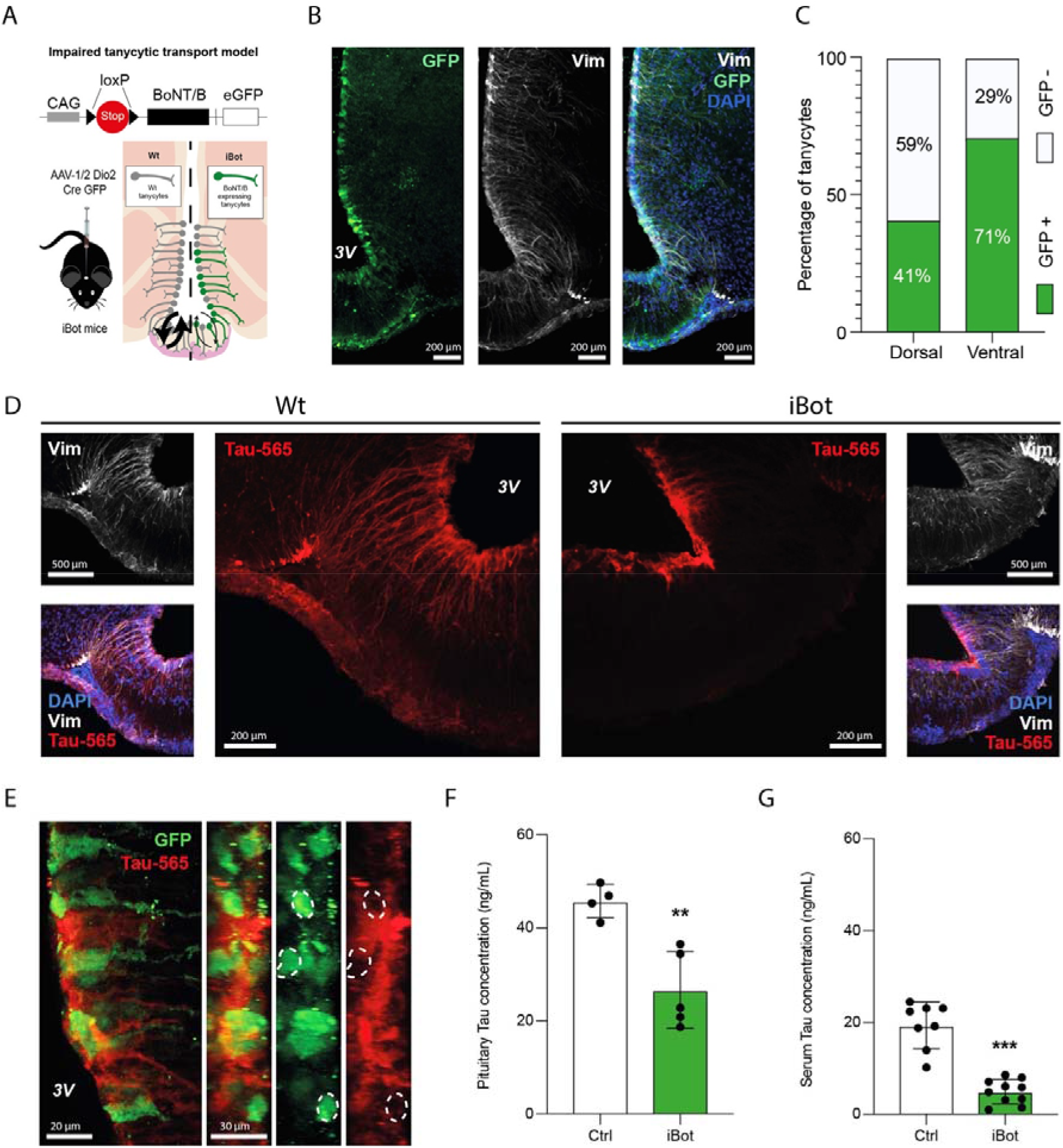
Impairing tanycytic vesicular transport blunts CSF-to-blood Tau transport. A. Schematic representation of the iBot mouse model. B. Photomicrograph of the median eminence of iBot mice injected with the AAV1/2-Dio2-Cre-A2-GFP virus, showing infected tanycytes (GFP, green) and immunolabeling for the tanycytic marker vimentin (white). C. Quantification of GFP-positive dorsal and ventral tanycytes in iBot mice injected with the AAV1/2-Dio2-Cre-A2-GFP virus (n=10 mice, 3-4 sections per mouse). D. Photomicrographs of Tau-565 (red) uptake in vimentin-immunopositive (white) tanycytes in the median eminence of Tau-565-injected WT mice (left panels) and iBot mice (right panels). E. Maximum intensity projection of the wall of the third ventricle of Tau-565 (red)-injected iBot mice, immunolabeled for GFP (green); z-stack acquisition (left panel) and z-axis reprojections (right panels) F. ELISA measurement of human Tau in the pituitary of Tau-565-injected WT and iBot mice 30 minutes post-injection (n= 4 and 5 mice). Two-tailed unpaired *t*-test (t_(11)_=4.293, p=0.004). Data are presented as means ± S.D., **, p<0.01. G. ELISA measurement of human Tau in the serum of Tau-565-injected WT and iBot mice 30 minutes post-injection (n=8 and 10 mice). Two-tailed unpaired *t*-test (t_(16)_=7.875, p<0.001). Data are presented as means ± S.D., ***, p<0.001.

### CSF-to-blood Tau efflux is age-dependent and altered in AD patients

Since CSF-to-blood ratios have been used in several studies to evaluate molecular transport into the brain across the BBB (*38, 39*), we considered the ratio of plasma to CSF Tau in AD patients to be a proxy for its clearance in the opposite direction. Indeed, although Tau protein concentrations have been previously compared and found to be higher in the CSF and plasma of AD patients compared to normal subjects (*40, 41*), the importance of the plasma-to-CSF Tau ratio as an indicator of Tau clearance from the brain has not been exploited so far. Plasma and CSF concentrations of total Tau protein for control and AD patients were extracted from the Alzheimer’s Disease Neuroimaging Initiative (ADNI1) database (*42*). After screening and data cleaning, 96 control patients and 88 AD patients, for whom plasma and CSF Tau concentrations were available at baseline, were selected. Mean plasma and CSF Tau levels were higher in AD patients (n=88, 3.128 ± 1.386 and 359 ± 131 pg/mL, respectively) than in control subjects (n=96, 2.503 ± 1.208 and 239 ± 85 pg/mL, p=0.001 and p<0.0001 respectively, Fig. 3A,B). Interestingly, however, the increase in Tau concentrations was not proportionate in the two compartments, and the plasma-to-CSF ratio of Tau was lower in AD patients (n=88, 0.01022 ± 0.005) than in controls (n=96, 0.01303 ± 0.007, p=0.005), suggestive of a defect in brain-to-blood Tau efflux in these patients (Fig. 3C). Remarkably, while the plasma-to-CSF Tau ratio was negatively correlated with age in control patients (r= −0.3378, p=0.0008, R^2^= 0.1141), indicative of a mild age-related decline in clearance, this correlation was lost in AD patients (r= −0.0008, p=0.99, R^2^= 6.717.10^−7^) (Fig. 3D; Supplementary Fig. 2A,B). However, when groups were stratified based on the median age of the cohort (75 years), the decrease in plasma-to-CSF Tau ratio values in AD patients compared to controls was evident in the younger but not in the older group, confirming the existence of an age-related decrease in clearance even in controls (Fig. 3E). Further analysis revealed that this age-related difference in the plasma-to-CSF Tau ratio was due to the fact that plasma Tau concentrations were not significantly lower in controls than in AD patients at younger ages, unlike older ones (Fig. 3F), while the increase in CSF Tau concentrations in AD patients compared to controls was greater in the younger group than in the older one, although both increases were significant (Fig. 3G).

**Figure 3:**
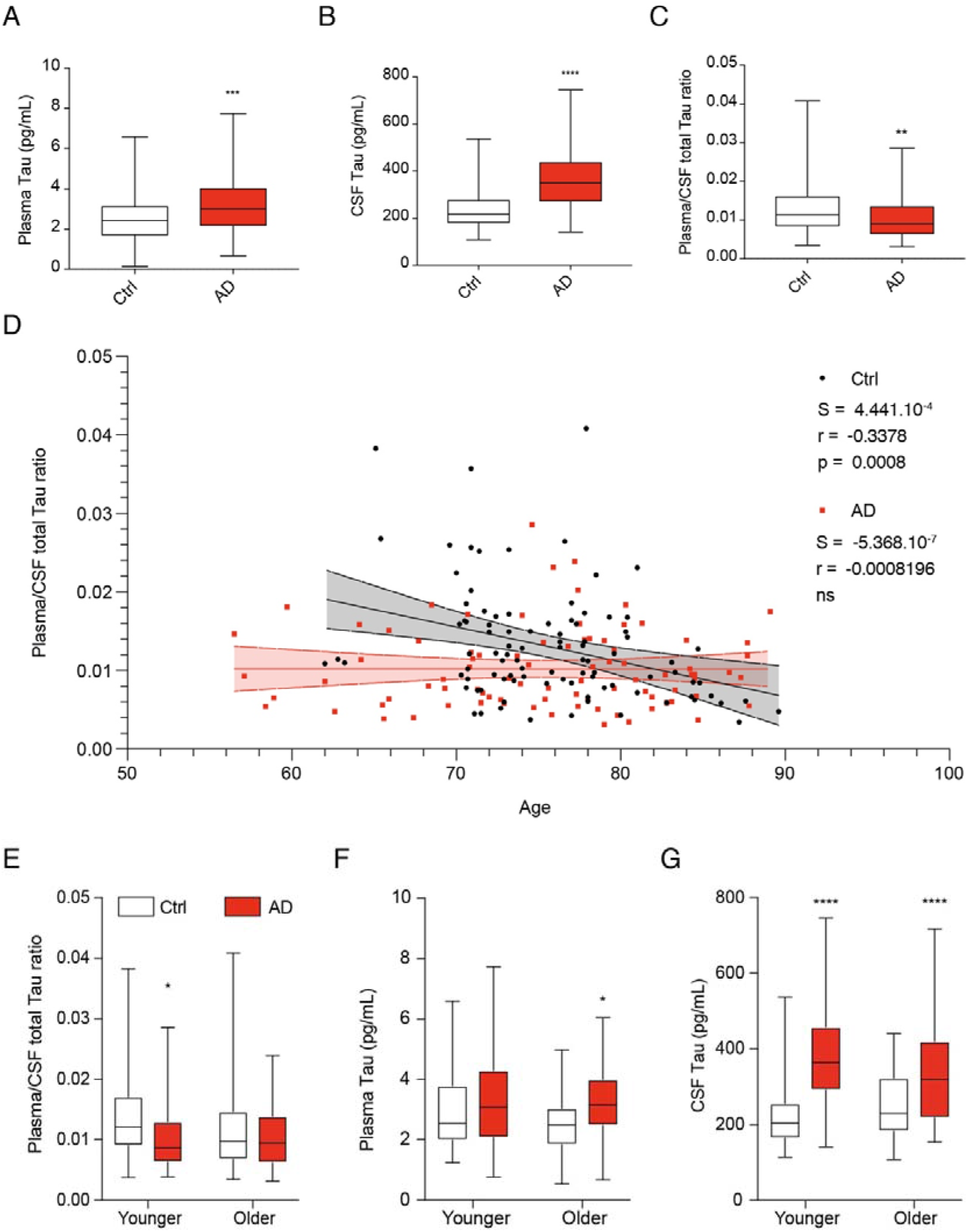
Plasma-to-CSF Tau ratio is lower in AD patients. A. Plasma Tau concentrations in control (white, n=96) and AD (red, n=88) patients. Two-tailed Mann-Whitney U test (*U*=3443, p=0.001). Data are presented as a box (median, lower and upper quartile) and whiskers (minimum and maximum); ***, p<0.001. B. CSF Tau concentrations in control (white, n=96) and AD (red, n=88) patients. Two-tailed Mann-Whitney U test (*U*=2062, p<0.0001). Data are presented as a box (median, lower and upper quartile) and whiskers (minimum and maximum); ****, p<0.0001. C. Plasma-to-CSF ratio of total Tau concentrations in control (white, n=96) and AD (red, n=88) patients. Two-tailed Mann-Whitney U test (*U*=3176, p=0.005). Data are presented as a box (median, lower and upper quartile) and whiskers (minimum and maximum); **, p<0.01. D. Plasma-to-CSF Tau ratio as a function of age in control (black) and AD (red) patients. Linear regression curve and 95% confidence interval for control (black dashed lines and grey shadow) and AD patients (red dashed lines and red shadow). E. Plasma-to-CSF Tau ratio stratified by age in control (younger: <75 years, n=49; older: >75 years, n=47) and AD (younger: <75 years, n=42; older: >75 years, n=46). Kruskal-Wallis test (*H*_(*4*)_=11.03, p=0.0116) followed by Dunn’s multiple comparison post hoc test (Younger Ctrl vs. Younger AD, *Q*_(4)_=2.995, p=0.0165). F. Plasma Tau concentrations stratified by age in control (younger: <75 years, n=49; older: >75 years, n=47) and AD (younger: <75 years, n=42; older: >75 years, n=46). Kruskal-Wallis test (*H*_(*4*)_=11.03, p=0.0103) followed by Dunn’s multiple comparison post hoc test (Older Ctrl vs. Older AD, *Q*_(4)_=2.679, p=0.0443). Data are presented as a box (median, lower and upper quartile) and whiskers (minimum and maximum); *, p<0.05. G. CSF Tau concentrations stratified by age in control (younger: <75 years, n=49; older: >75 years, n=47) and AD (younger: <75 years, n=42; older: >75 years, n=46). Kruskal-Wallis test (*H*_(*4*)_=53.64, p<0.0001) followed by Dunn’s multiple comparison post hoc test (Younger Ctrl vs. Younger AD, *Q*_(4)_=6.465, p<0.0001; Older Ctrl vs. Older AD, *Q*_(4)_=3.430, p<0.0001). Data are presented as a box (median, lower and upper quartile) and whiskers (minimum and maximum); ****, p<0.0001.

### Tanycytes transport Tau in the human brain but are degraded in AD

Given our experiments above in rat primary tanycytes *in vitro* and mice *in vivo* and the finding that Tau efflux from the CSF to the blood is altered in AD patients, we used human post mortem brain tissues from six AD patients and five controls (Table 1) to further examine the hypothesis that tanycytes are Tau transporters. Using immunofluorescence labeling with the PHF1 antibody (anti-Tau pSer396/404), we noted no signal in the brain of control subjects (Fig. 4A-B) but Tau-immunoreactive neurons with typical neurofibrillary tangles in the infundibulum of AD patients (Fig. 4B). Interestingly, tanycytes displayed a pattern of Tau labeling that was quite different from neurofibrillary tangles, with Tau-positive punctae corresponding to vesicles both at the level of their cell bodies lining the third ventricle and along the proximal part of their vimentin-immunoreactive processes (Fig. 4B). This distinctive pattern, similar to our observations with Tau-565 in rat tanycytes, suggests that the immunolabeling may correspond to Tau endocytosed from the CSF, rather than to Tau naturally produced by tanycytes. Strikingly, however, the morphology of tanycytes in AD patient brains was dramatically different from that in controls. Indeed, their vimentin cytoskeleton appeared fragmented, their processes seemed to be discontinuous and an accumulation of small vimentin-positive bodies could be observed (Fig. 5A). To better understand the changes occurring in AD patient brains, we used the Ilastik segmentation toolkit to perform a machine-learning-aided analysis of vimentin immunolabeling (*43*). After pixel classification, the detected objects were classified into two groups: “processes”, consisting of long, slim objects corresponding to tanycytic projections, and “fragments”, consisting of small rounded objects corresponding to putative tanycytic debris or truncated projections (Fig. 5B). This analysis suggested that intact tanycytic processes were much fewer in number and shorter in length in AD patient brains than in controls, and conversely, that fragmented or damaged tanycytic processes were more numerous and of larger diameter in AD patients (Supplementary Fig. 3A-D). In agreement with this observation, the area covered by “processes” was lower in AD patients compared to controls, whereas the area covered by “fragments” was higher in AD patients than in controls (Fig 5C,D). Furthermore, in control subjects, 95% of the area covered by vimentin immunolabeling corresponded to tanycytic “processes”, whereas these represented only 57% of the vimentin signal in AD patients (n=5 for each group, p<0.001, Fig. 5E), highlighting a profound degradation of the structure of tanycytes in AD.

**Table 1:** Patients information.

|  | Sex | Group | Age | Braak stage | Thal stage | Post-mortem delay | Comorbidities | PHF1 signal Hypothalamus | Tanycytic degradation |
| --- | --- | --- | --- | --- | --- | --- | --- | --- | --- |
| Patient 1 | M | Ctrl | 50's | 0 | 0 | 16h | Ischemic cardiopathy, epilepsy, dyslipidemia | 0 | 0 |
| Patient 2 | F | Ctrl | 60's | 0 | 0 | 27h | Arterial hypertension, atrial fibrillation, aortic infectious endocarditis, strokes | 0 | 0 |
| Patient 3 | M | Ctrl | 30's | 0 | 0 | 80h | Atherosclerosis, obesity | 0 | 0 |
| Patient 4 | M | Ctrl | 60's | 0 | 0 | 19h | Stroke, arterial hypertension, dyslipidemia | 0 | 0 |
| Patient 5 | M | Ctrl | 70's | I | 0 | 21h | obliterative arteriopathy of the lower limbs, chronic glaucoma | 0 | 0 |
| Patient 6 | F | AD | 80's | IV | IV | 27h | Breast cancer, arterial hypertension | +++ | +++ |
| Patient 7 | M | AD | 50's | VI | IV | 32h | No history | +++ | +++ |
| Patient 8 | M | AD | 50's | IV | V | 14h | Smoking | +++ | ++ |
| Patient 9 | F | AD | 50's | V/VI | IV | 36h | arterial hypertension, atrial fibrillation, infectious endocarditis | 0 | +++ |
| Patient 10 | F | AD | 90's | III | III | 24h | Horton disease, arterial hypertension, dysthyroidism, osteoporosis, obstructive sleep apnea | 0 | +++ |
| Patient 11 | M | AD | 70's | V/VI | II | 48h | Arterial hypertension | ++ | ++ |
| Patient 12 | F | FTD Tau | 60's | NA | NA | 40h | Asthma | ++ | 0 |
| Patient 13 | M | FTD TDP-43 | 70's | NA | NA | 24h | No history | 0 | 0 |
| Patient 14 | F | FTD TDP-43 | 70's | IV | 0 | 20h | AD associated Tau lesions but absence of amyloid beta | + | ++ |
| Patient 15 | M | FTD TDP-43 | 70's | NA | NA | 12h | Presence of some Tau lesions in astrocytes, dyslipidemia | ++ | 0 |

**Figure 4:**
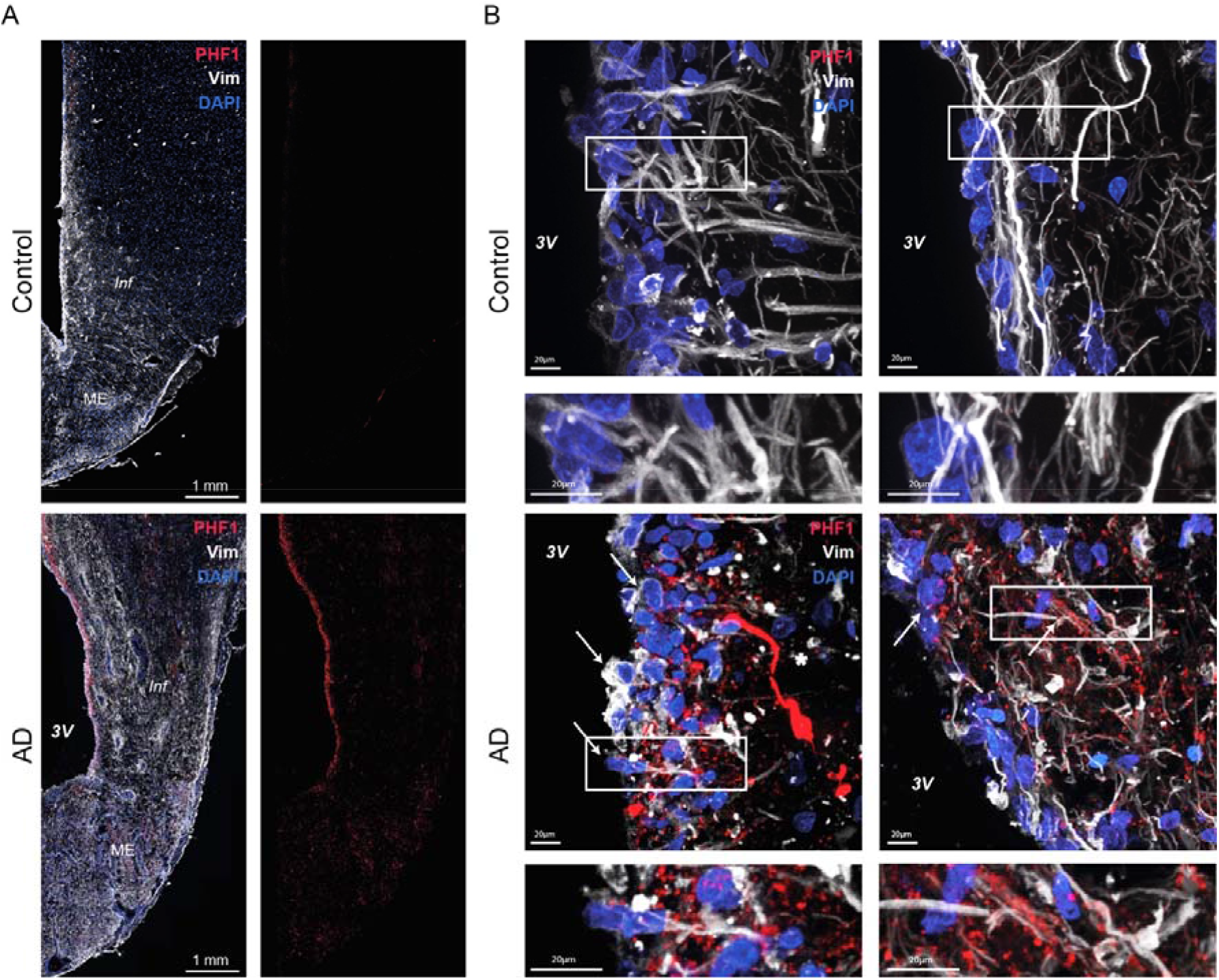
Tanycytes of AD patients contain Tau-positive vesicles. A. Photomicrograph of tanycytes in the hypothalamus of control and AD patients immunolabeled for vimentin (white) and phosphorylated Tau (PHF1 antibody, Tau pSer396/404, red). B. Higher magnifications of tanycytic cell bodies and processes in the control and AD patients brain, showing a typical neuron with a neurofibrillary tangle labeled by PHF1, and more punctate labeling corresponding to Tau-containing vesicles in tanycytes only in AD patients. White rectangle: higher magnification inset shown at right. White arrows: tanycytes containing PHF1-positive vesicles, white asterik: PHF1-positive neurofibrillary tangle.

**Figure 5:**
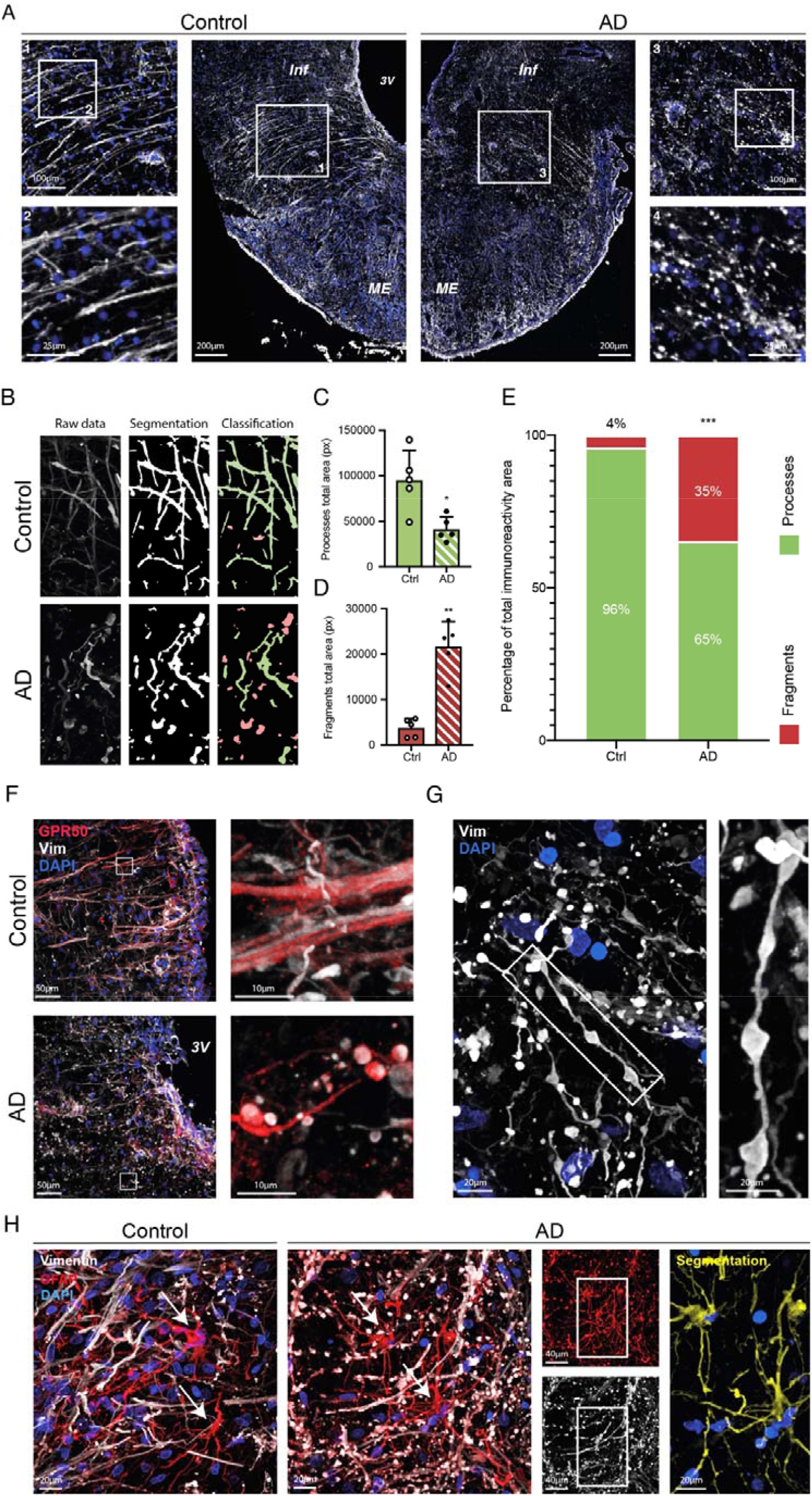
Tanycytes in AD patients are degraded and disorganized but not astrocytes. A. Photomicrographs of tanycytes in the median eminence and infundibulum of control (left panels) and AD patients (right panels) immunolabeled for vimentin (white). Insets: higher magnification images of tanycytic processes in control (left panels) and AD patients (right panels), showing fragmentation of vimentin-positive processes in the latter. B. Representation of the Ilastik analysis pipeline starting from raw data acquisition of vimentin immunolabeling (left panels) to segmentation of the vimentin signal (middle panels) to classification of tanycytic processes and fragments (green and red respectively, right panels). C. Graph representing the total area covered by processes in control (n=5) and AD patients (n=5). Two-tailed Mann-Whitney U test (*U*=1, p=0.02). Data are presented as means ± S.D. *, p<0.05. D. Graph representing the total area covered by fragments in control (n=5) and AD patients (n=5). Two-tailed Mann-Whitney U test (*U*=0, p=0.008). Data are presented as means ± S.D. **, p<0.01. E. Area covered by processes (green) vs. fragments (red) as a percentage of the total vimentin immunoreactive area in control (n=5) and AD patients (n=5). Two-way ANOVA (patient condition, *F*_(1,16)_=2.125.10^−20^, p>0.99; object class, *F*_(1,16)_=696.2, p<0.001; interaction, *F*_(1,16)_=173.1, p<0.001) followed by Sidak’s multiple comparison post hoc test (Ctrl Processes vs. Ctrl Fragments, *q*_(16)_=27.96, p<0.001; AD Processes vs. AD Fragments, *q*_(16)_=9.355, p<0.001; Ctrl Fragments vs. AD Fragments, *q*_(16)_=9.302, p<0.001; Ctrl processes vs. AD processes, *q*_(16)_=9.302, p<0.001). Data are presented as means; ***, p<0.001. F. Photomicrographs of tanycytes in the median eminence of control (top panels) and AD patients (bottom panels) immunolabeled for vimentin (white) and GPR50, a tanycytic membrane protein (red). Insets: high magnification photomicrographs of tanycytic processes in control (top panel) and AD patients (bottom panel) showing that the fragmentation in AD patients involves the entire tanycytic process and not just the vimentin cytoskeleton. G. Photomicrographs of tanycytes in the median eminence of an AD patient immunolabeled for vimentin (white). Inset: high magnification showing the “string of pearls” appearance of tanycytes in AD patients. H. Photomicrographs of the median eminence of control (left panel) and AD patients (right panels) immunolabeled for vimentin (white; tanycytes) and GFAP (red; tanycytes and astrocytes). The rightmost panel is a segmentation for astrocytes (GFAP not overlapping with vimentin in yellow, DAPI in blue). Arrows indicate astrocytic cell bodies. Inset: region subjected to segmentation at right.

Next, to verify whether this degradation was limited to the vimentin cytoskeleton or whether the structural integrity of the entire cell was affected, we co-immunolabeled tissues for vimentin as well as GFAP, another cytoskeletal protein that interacts with vimentin in tanycytes to form the secondary cytoskeleton, or GPR50, a membrane receptor whose expression in the infundibulum is limited to tanycytes (*44*). GFAP immunolabeling was seen in tanycytic structures in both control subjects and AD patients, most often colocalizing with vimentin, and reproduced the morphological alterations observed with the latter (Supplementary Fig. 3E). Similarly, GPR50, which was also expressed by tanycytes in both controls and AD patients, appeared to envelop vimentin-positive “fragments” rather than delineating intact processes in AD patients (Fig. 5F). Interestingly, however, in some instances, fine vimentin-positive filaments could be seen linking the putative fragments of tanycytic process, giving rise to the appearance of a “string of pearls” (Fig. 5G), indicating a condensation or aggregation of the tanycytic cytoskeleton or cytoplasmic contents to form bead-like structures rather than the disintegration of the cell or process as a whole. Strikingly, while tanycytic processes positive for both vimentin and GFAP presented a fragmented appearance, astrocytes expressing GFAP, but not vimentin, in the vicinity of these tanycytes did not display similar alterations in AD patients, suggesting that the degradation observed was specific to tanycytes (Figure 5H), and moreover, not an artifact caused by the plane of sectioning. As might be expected, this degradation of tanycytic processes also disrupted tanycyte-capillary interactions in AD patient brains, as seen using double immunolabeling for vimentin to identify tanycytic endfeet and caveolin 1 (Cav1) to label capillary walls (Supplementary Fig. 3F), implying functional consequences for blood-brain exchanges.

Given the 3D distribution of Tau-transporting tanycytes in the mouse brain, we then wondered whether the degradation of tanycytes in the brain of AD patients was uniform or spatially differentiated. Interestingly, although an examination of vimentin immunolabeling in sagittal sections of the ME showed that these changes occurred all along the anteroposterior axis of the ME (Fig. 6A), the putative fragmentation or beading of the tanycytic cytoskeleton in all patients was more pronounced in the infundibulum and the internal zone of the ME, close to the third ventricle, but was observed less frequently or not at all in the external part of the ME (Fig. 6B). Finally, to determine whether this tanycytic degradation was specific to AD or could be more generalized, we compared vimentin immunolabeling in the brain of control subjects, AD patients and patients with frontotemporal dementia (FTD) of either the TDP-43 or Tau type. Intriguingly, regardless of FTD type, the tanycytic processes of FTD patients (n=4) were morphologically different from those in controls; however, except in one FTD patient brain that also revealed signs of AD post mortem (see Table 1), tanycytes did not display any beading (Fig. 6C). Furthermore, Ilastik-based analysis of vimentin immunolabeling showed that FTD patients had a lower density of “processes” than control subjects, similar to AD patients (Fig. 6D). However, the reduced “process” area in FTD was not accompanied by an increase in the area covered by “fragments”, unlike in AD patients (Fig. 6E). In addition, the proportion of “processes” to “fragments” was similar to that in controls, supporting a degradation of the tanycytic network without their disintegration.

**Figure 6:**
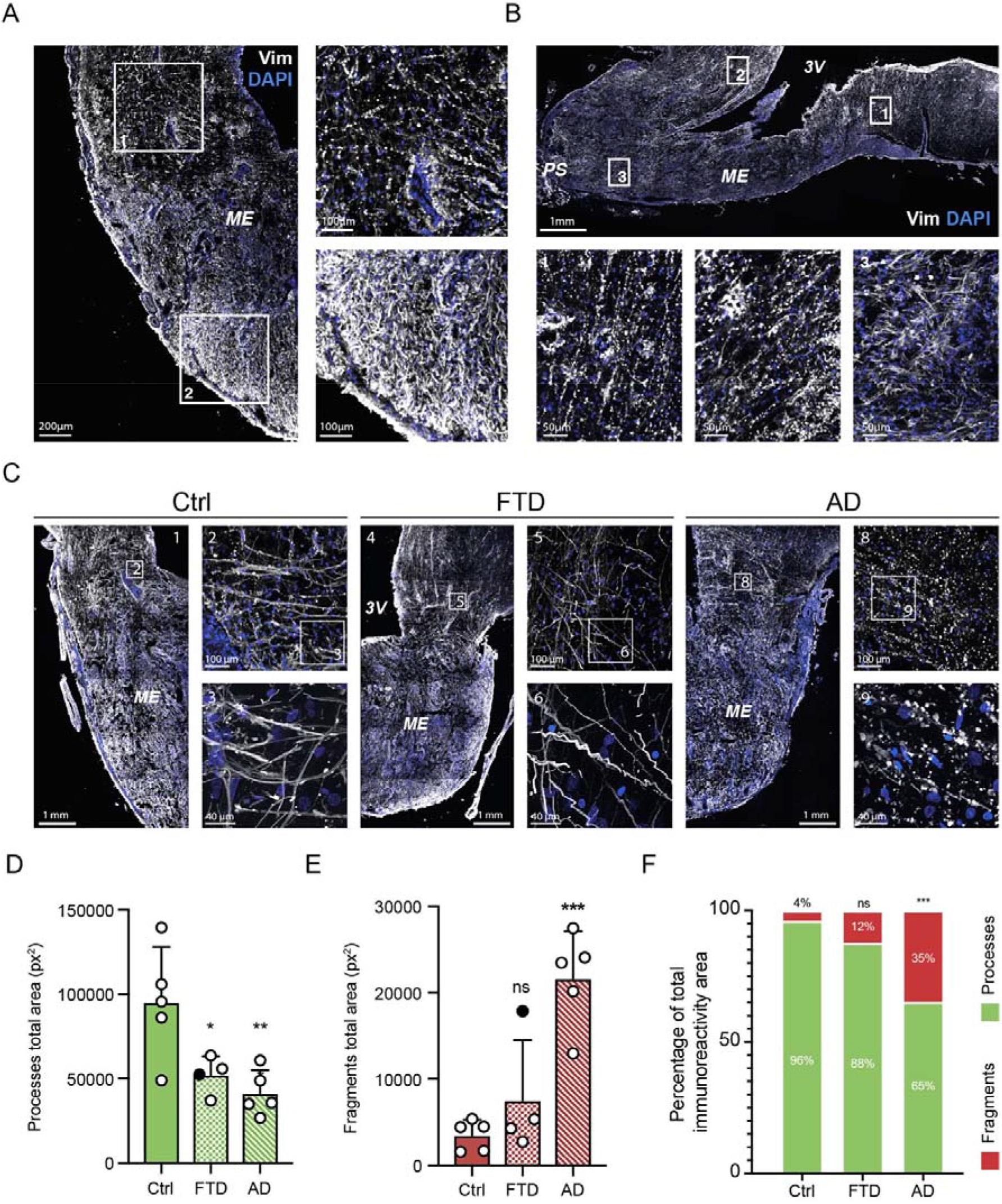
Tanycytic degradation occurs throughout the median eminence in AD but not in FTD. A. Photomicrographs of a median eminence in coronal section from an AD patient showing tanycytes immunolabeled for vimentin (white). Inset 1: higher magnification of the infundibular region. Inset 2: higher magnification of the external median eminence region. B. Photomicrograph of a median eminence in sagittal section from an AD patient showing tanycytes immunolabeled for vimentin (white). Inset 1: higher magnification of the posterior region of the median eminence. Inset 2: higher magnification of the median eminence adjoining the optic chiasma. Inset 3: higher magnification of the external median eminence close to the pituitary stalk. C. Photomicrographs of the median eminence and infundibulum of control (left panels), FTD (middle panels) and AD patients (right panels) showing tanycytes immunolabeled for vimentin (white). Numbered insets: localization of higher magnification photomicrographs showing the difference in tanycytic morphology between the three conditions. D. Total area covered by tanycytic processes in control (n=5), FTD (n=4) and AD patients (n=5). One-way ANOVA (F_(2,11)_=8.136, p=0.0068) followed by Tukey’s multiple comparison post-hoc test (Ctrl vs. AD, q_(11)_=5.433, p=0.0071; Ctrl vs. FTD, q_(11)_=4.096, p=0.0358; AD vs. FTD, q_(11)_=1.026, p=0.7538). Data are presented as means ± S.D. **, p<0.01; *, p<0.05. The FTD patient represented by a black dot also showed signs of AD in the brain post mortem. E. Total area covered by tanycytic fragments in control (n=5), FTD (n=4) and AD patients (n=5). One-way ANOVA (F_(2,11)_=17.02, p=0.0004) followed by Tukey’s multiple comparison post-hoc test (Ctrl vs. AD, q_(11)_=7.884, p=0.0004; Ctrl vs. FTD, q_(11)_=1.572, p=0.5269; AD vs. FTD, q_(11)_=5.861, p=0.0043). Data are presented as means ± S.D. ***, p<0.001; ns, non-significant. The FTD patient represented by a black dot also showed signs of AD in the brain post mortem. F. Area covered by tanycytic processes (green) and fragments (red) as a percentage of the total vimentin immunoreactive area in control (n=5), FTD (n=4) and AD patients (n=5). Two-way ANOVA (patient condition, *F*_(2,22)_=3.280.10^−18^, p>0.99; object class, *F*_(2,22)_=687.2, p<0.001; interaction, *F*_(2,22)_=58.39, p<0.001) followed by Sidak’s multiple comparison post hoc test (for processes: Ctrl vs. FTD, *q*_(22)_=1.791, p=0,2391; Ctrl vs. AD, *q*_(22)_=7.276, p<0.0001; FTD vs. AD, *q*_(22)_=5.069, p<0.001; for fragments: Ctrl vs. FTD, *q*_(22)_=1.791, p=0,2391; Ctrl vs. AD, *q*_(22)_=7.276, p<0.0001; FTD vs. AD, *q*_(22)_=5.069, p<0.001).Data are presented as means; ***, p<0.001.

Together, these observations support the hypothesis that tanycytic pathology could be disease-specific, and the degradation of the tanycytic cytoskeleton in AD patients likely interferes with Tau efflux from the CSF, contributing to its accumulation in the brain.

## DISCUSSION

The accumulation and aggregation of both Aβ and Tau are hallmarks of AD and contribute to AD pathogenesis. Aβ clearance has been widely studied and involves both intra- and extracellular degradation, blood-brain barrier crossing, interstitial fluid clearance and cerebrospinal fluid (CSF) clearance by absorption (*45, 46*). On the other hand, while Tau degradation is well documented (reviewed in (*47*)), the mechanisms underlying its transport out of the brain and the exact paths that it follows to cross the BBB and reach the peripheral circulation are still poorly understood. Our findings above indicate that tanycytes are a significant contributor to Tau efflux from the CSF to the blood and that this clearance pathway might be impaired in the context of AD due to the morphological alteration of tanycytic processes, and presumably, the interruption of vesicular transport. Furthermore, our observations in FTD patient brains suggest that tanycytic degradation or dysfunction may be a more widespread feature of neurodegenerative disorders than currently imagined.

Several studies have used the injection of different Tau isoforms into the brain to highlight its ability to exit the parenchyma and/or cross the BBB (*13, 48*), and have proposed the involvement of various brain structures such as the choroid plexus, arachnoid villi and a brain lymphatic system in this process (*11, 49*). In our model of Tau ICV injection, the choroid plexus did not show any signs of Tau uptake. Additionally, while the contribution of the lymphatic system and arachnoid villi could not be assessed due to the destruction of the meninges during brain dissection, another study has revealed only a moderate effect of the disruption of the brain lymphatic system on brain-to-blood Tau transfer (*33*). On the other hand, in our study, specifically disrupting vesicular transport in a significant proportion of tanycytes, but not all of them, resulted in an incomplete but robust inhibition of Tau efflux to the periphery that lymphatic transport could not adequately compensate for. While the contribution of other players and processes such as BBB degradation and leakage cannot be ruled out, our experimental paradigm still reveals a major role for tanycytes in Tau efflux.

While Tau accumulation in the brain in the context of AD has been observed in numerous studies, from mouse models to AD patients, studies describing circulating Tau levels in mice are rare (*14, 19, 48, 50*). The existence of an AD patient database that includes Tau concentrations in the CSF and blood allowed us to indirectly evaluate Tau efflux by calculating the relative concentrations of Tau in the destination tissue, i.e. blood or plasma, vs. the compartment of origin, i.e. the CSF, as is often done in the reverse direction for other molecules. Although this value represents the balance of all influx/efflux mechanisms and is not specific to any single transport system, it is the only readout of Tau transport available in live patients. Interestingly, CSF concentrations of Tau in the AD group were already high at younger ages, whereas they gradually increased with age in control patients, catching up with the AD group. The resulting increase in Tau plasma-to-CSF ratio in younger AD patients sets this process apart from the physiological aging seen in control subjects. Age-related changes, including cell loss and impaired signaling cascades, have previously been observed in the tanycytes of both women and male rats (*51, 52*). Although not previously noticed, our observation of tanycytic degradation across AD patients at different ages and stages of disease (our post mortem brains were obtained from patients ranging from 52-88 years old, but at Braak stages III-IV; Table 1), suggests that these changes could accompany or even contribute to increased Tau accumulation in the brain by disrupting its transport, rather than being secondary to Tau accumulation. On the other hand, in older AD patients, a compensatory efflux mechanism such as BBB leakage could explain why Tau does not continue to accumulate in the CSF, and why plasma Tau concentrations are also significantly higher than in age-matched controls.

The progressive “beading” of distal processes, a phenomenon known as clasmatodendrosis, has long been described to occur in astrocytes under a variety of pathological conditions (*53–55*). In addition, amyloid plaques in animal models appear to trigger the phenomenon in surrounding astrocytes (*56, 57*), and age-related astrocytic clasmatodendrosis is associated with an accumulation of Aβ in the hippocampus (*58*). However, astrocytes in our patient brain sections displayed a perfectly normal morphology. In addition, there are few reports of the process in human AD brains, and tanycytic clasmatodendrosis, or for that matter, any other morphological alteration in tanycytes, has not so far been described either in AD or in any other human disorder, and its causes, mechanisms and consequences deserve to be further explored.

Regardless of the lack of prior evidence of tanycytic structural abnormalities, several disorders have been shown to be associated with their dysfunction. Notably, preclinical studies have revealed that impaired tanycytic function could underlie metabolic diseases such as type 2 diabetes and obesity, by interfering with the access of peripheral signals to hypothalamic circuits regulating metabolism and energy homeostasis (*28, 30–32*). Intriguingly, these conditions are also associated with an increased risk of developing AD, suggesting that tanycytes could be the long-sought missing link that is thought to exist between metabolic disorders and AD (*27, 59*). However, the clinical manifestations of AD extend beyond cognitive and metabolic deficits, and include several features that could be attributed to neuroendocrine dysfunctions (*60*). In this context, ME tanycytic endfeet also interact with the terminals of neuroendocrine neurons to regulate the secretion of neurohormones of the hypothalamic-pituitary axes (*27*), suggesting that tanycytic pathology could also lie at the root of hormonal disturbances in AD, which themselves could also contribute to age-or disease-related cognitive and metabolic deficits (*60*).

Finally, the fact that tanycytes are able to transport Tau and their very different alteration in FTD raises broader questions as to their role in the development of neurodegenerative diseases. The fact that they did not transport fluorescent BSA, which has a similar molecular weight, together with the observation of endogenous Tau in tanycytic vesicles in human AD brains, is significant, as it suggests that there might be some selectivity in terms of their cargo, but could they also transport other pathological molecules such as Aβ or alpha-synuclein? How does the fact that they are morphologically different in FTD as compared to both controls and AD patients, as well as sparser or thinner in the former, affect their diverse functions? In addition, the directionality of their transport of Tau, from the CSF to the blood circulation, is itself striking, since these cells have until now been associated with the transcytotic shuttling of blood-borne molecules into the CSF. In effect, this two-way secretory activity of tanycytes – allowing meaningful metabolic hormones and signals to be captured from the circulation and released into the brain, while performing the reverse function for a molecule that needs to be eliminated, such as Tau, without themselves producing either of the two types of molecules released – corresponds to neither conventional secretion nor excretion. An apt term for such a sufficiently singular phenomenon could be “metacrine” secretion, to indicate a process that surpasses mere secretion. Exploring the molecules and mechanisms involved in this phenomenon and its involvement in normal physiology and pathological processes could represent a new avenue of research in the interaction of the brain and the periphery.

## MATERIALS AND METHODS

### Collection and processing of human tissues

Tissues were obtained in accordance with French laws (Good Practice Concerning the Conservation, Transformation and Transportation of Human Tissue to be Used Therapeutically, published on December 29, 1998). Permission to use human tissues was obtained from the French Agency for Biomedical Research (Agence de la Biomedecine, Saint-Denis la Plaine, France, protocol no. PFS16-002) and the Lille Neurobiobank. Ethics committee approval for this work was provided by the Comité de Protection des Personnes (CPP) SUD-EST II (BIOWATCH, #2021-A00879-32).

Dissected blocks of the adult brain containing hypothalamus were fixed by immersion in 4% paraformaldehyde in PBS, pH 7.4 at 4°C for 1 week. The tissues were cryoprotected in 30% sucrose/PBS at 4°C until the fragment sunk, embedded in Tissue-Tek OCT compound (Sakura Finetek), frozen in dry ice and stored at −80°C until sectioning. Fragments were cut either in coronal or sagittal sections of 20 μm.

### Immunohistology

For human hypothalamus immunolabeling, a citrate-buffer antigen retrieval step, 10mM Citrate in TBS-Triton 0.1% pH 6 for 30 min at 70°C, was performed on 20μm sections. For mice brain immunolabeling, no antigen retrieval step was performed. After 3 washes of 5 minutes with TBS-Triton 0.1%, sections were blocked in incubation solution (ICS: 10% normal donkey serum, 1mg/ml BSA in TBS-Triton 0.1% pH 7,4) for 1 hour. Blocking was followed with primary antibody incubation (Supplementary Table 1) in ICS for 48h at 4°C. Primary antibodies were then rinsed out, before incubation in fluorophore-coupled secondary antibodies for 1h in ICS at room temperature. Secondary antibodies were washed and sections counterstained with DAPI (D9542, Sigma). Finally, sections were treated with Autofluorescence Eliminator Reagent (2610, Merck Millipore) to quench lipofuscin aggregates autofluorescence before mounting using Mowiol.

### Ilastik segmentation analysis

Ilastik toolkit (*43*) was used for tanycytic processes and fragments segmentation analysis. We used the pixel classification followed by object classification pipelines to segment vimentin signal and classify vimentin positive objects into two classes: the “process” class including long and slim vimentin positive tanycytic processes and the “fragment” group including short and circular vimentin positive tanycytic fragments. To this end, high magnification z-stack confocal acquisitions (x63, 80 stacks, 0,25μm z-step) of vimentin immunolabeled hypothalami were acquired for all patients and transformed in maximum intensity projections. First, a set pictures of control and AD patients (5 patients per group, 1 section per patient, 3 images per section) was used to train a pixel classification and an object classification algorithm based on user’s inputs. Then, a second set of control and AD patients’ acquisitions (5 patients per group, 5 sections per patient, 2-3 pictures per sections) were analyzed using the trained pipelines. Finally, the data was extracted and compiled as mean per patients and mean per groups. The relative coverage of processes and fragments was computed as the percentage of processes area or fragments area over the total immunoreactive area, being the sum of processes area and fragments area.

### ADNI data extraction and analysis

Data for control and AD patients were extracted from patients included in the ADNI1 cohort and for which CSF and plasma Tau concentrations at baseline were available (from ADNIMERGE, version: 2013-04-29 for baseline CSF Tau and BLENNOWPLASMATAU, version: 2015-08-04, for baseline plasma Tau). Plasma tau was analyzed by the Single Molecule array (Simoa) technique and the Human total tau assay that uses a combination of monoclonal antibodies that give a measure of total tau levels. CSF Tau was measured using the Research Use Only (RUO) INNOBIA AlzBio3 immunoassay (Fujirebio).The ADNI was launched in2003 as a public-private partnership, led by Principal Investigator Michael W. Weiner, MD. The primary goal of ADNI has been to test whether serial magnetic resonance imaging (MRI), positron emission tomography (PET), other biological markers, and clinical and neuropsychological assessment can be combined to measure the progression of mild cognitive impairment (MCI) and early Alzheimer’s disease (AD). A total of 105 control patients and 95 AD patients were found in the database. From CSF and plasma Tau concentration, CSF to plasma Tau ratio was calculated for each patient. Patients identified as outliers for CSF, plasma or CSF to plasma Tau by the ROUT method (Q=0,1%) were excluded resulting in the inclusion of 96 control and 88 AD patients for final analysis. To stratify patients into two groups of age (younger and older group) the median age of the cohort (75,8 years-old) was used (younger, n= 49 and 42; older, n= 47 and 46; for control and AD patients respectively).

### Animals

All C57Bl/6J adult male mice were housed under specific pathogen-free conditions in a temperature-controlled room (21–22□°C) with a 12-h light/dark cycle and 40% humidity, and ad libitum access to food and water. All experiments were performed on 2 to 4 months old mice. B6.FVB-Tg(CAG-boNT/B,-EGFP)U75-56Fwp/J (iBot) mice (JAX:#018056) were purchased from *Jackson Laboratories* (*35*). All experiments and procedures involved in this study were approved by the Ethics Committee of the Universities of Lille, in accordance with European Union norms for animal experimentation.

### Tau-565 and BSA-565 production

2N4R recombinant human Tau protein was produced as previously described (*61*). Tau protein and purified bovine serum albumin (BSA, A7030, Sigma Aldrich) were labeled with Atto-565-NHS-ester (72464, Sigma Aldrich) using a 4-fold molar excess of Atto-565-NHS-ester at 4°C for 4 hours. After labelling, 15 mM of Tris was added to quench the reaction and the proteins were centrifugated in Zeba desalting columns (87767, ThermoFisher Scientific) to remove any unreacted fluorophores. To ensure protein coupling, a mass spectrometry analysis of the conjugated Tau protein (Tau-565) was performed using a Shimadzu AXIMA Assurance Linear MALDI-TOF Mass Spectrometer showing the coupling of a maximum of 5 Atto-565 label per protein. Finally, average label incorporation was determined by measuring fluorescence and protein concentration (A_max_ x MW of protein / [protein] x ε_dye_). Tau-565 and BSA-565 label incorporation was respectively estimated at around 2 moles and 3 moles/mole of proteins.

### Tau-565 and AAV1/2-Dio2-iCre-A2-GFP delivery

AAV1/2-Dio2-iCre-A2-GFP (1,25.10^9^ genomic particles per μL) was produced as detailed previously (*62*). Tau-565 was produced as detailed above and adjusted to a 1 μg.μL^−1^ concentration before injection. Both AAV1/2-Dio2-iCre-A2-GFP and Tau-565 were stereotaxically infused into the lateral ventricle (1μL over 5 minutes, anteroposterior: −0,3mm, midline: ± 1mm, dorsoventral: −2,5mm) of wild-type (wt) or transgenic iBot isofluorane-anesthesized mice. For dual injections, wt and iBot mice were injected with AAV1/2-Dio2-iCre-A2-GFP three weeks before Tau-565 injections.

### Evaluation of AAV1/2-Dio2-iCre-A2-GFP infection efficiency

Three weeks after LV AAV1/2-Dio2-iCre-A2-GFPinfusion in iBot mice, animals were sacrificed, their brain collected and post-fixed by 4% paraformaldehyde in PBS (PFA) immersion overnight. Brains were cryoprotected in 30% sucrose overnight, embedded in TissuTek (Sakura) and frozen. Coronal sections (40μm) were cut and processed for immunofluorescence using chicken anti-vimentin (1:1,000; PCK-594P, BioLegend) and rabbit anti-GFP (1:5,000; A-11122, Invitrogen) primary antibodies revealed with donkey anti-chicken Alexa Fluor 647 (1:1,000; 703-545-155; Jackson Immuno Research) and donkey anti-rabbit Alexa Fluor 488 (1:1000; A-21206, ThermoFisher Scientific). Sections were counterstained with DAPI and mounted using mowiol. Images were acquired using (spinning disk spec). Five representative ME slides per mice were analysed and tanycytes were divided into two groups (ventral for tanycytes projecting to the ME and dorsal for tanycytes projecting in the hypothalamic nuclei). Vimentin^+^/GFP^+^ and vimentin^+^/GFP^−^ cell number was reported to vimentin^+^ cells bordering the third ventricle.

### Tau-565 kinetic experiment

For experiments assessing Tau-565 clearance from brain to blood, anesthetized wt mice were stereotaxically injected with 1μL of Tau-565 (1μg.μL^−1^) over 5 minutes in the lateral ventricle. After 15, 30 minutes, 1- and 2-hours brains, pituitaries and blood of the mice were collected (n=5 per group, except for the 30 minutes group n=3). The brains were immersion fixed overnight at 4°C in 4% PFA. The pituitaries were immediately frozen in dry-ice and serum prepared from blood by centrifugation (2,000g, 15 min at 4°C).

### IDISCO tissue clearing

Immersion fixed brains were processed using an adapted version of the iDISCO+ protocol described previously (*63*). Briefly, samples were dehydrated with ethanol gradient (20%, 40%, 60%, 80%, 100%, 1 hour each) and delipidated in 66% dichloromethane / 33 % ethanol overnight. Ethanol was washed out of the samples by 100% dichloromethane incubation for 1h. Finally, the samples were cleared by immersion in dibenzylether for at least 2 hours in rotation. After transparency was achieved, a fresh solution of dibenzylether was used for storage, and the samples were kept protected from light at room temperature until imaging.

### Light-sheet microscope imaging

Imaging of cleared tissues was performed in dibenzylether on the Ultramicroscope 1 (Lavision BioTec, available at the BioImaging Center of Lille) and using MI PLAN 12x/0.53 with 2x zoom objective. Sequences were acquired with ImspectorPro Software. The following parameters were used: z-step was set to 2 μm, laser width and numerical aperture were kept to the maximum.

### 3D image processing and analysis

Tiff sequences resulting from light-sheet acquisitions were converted to the Imaris file format using Imaris FileConverter. Finally, Imaris 9.1-9.5 (Bitplane) was used for visualization and 3D processing of the datasets. Figures were prepared in Photoshop (Adobe) and videos were edited with Shotcut (https://shotcut.org/).

### Human Tau ELISA

For human Tau ELISA, pituitary protein extract was prepared by mechanical tissue dissociation in 1x RIPA buffer (20-188, Millipore) using tube potters and sonication. For primary tanycyte Tau-565 secretion experiment, cell medium was collected and immediately frozen in dry-ice. Tau-565 concentrations in pituitary extracts, serum and cell medium were measured using Tau (Total) Human ELISA Kit (KHB0042, ThermoFisher Scientific) according to manufacturer instructions except for 1:10 pituitary extract (0,2μg.μL^−1^), serum and cell medium dilution.

### Tanycyte primary culture

Tanycyte were isolated from the median eminence (ME) of the hypothalamus of 10-days-old rats and cultured as described previously (*64*). Briefly, after decapitation and removal of the brain, MEs were dissected and dissociated using a 40μm nylon mesh. Dissociated cells were culture in DMEM without pyruvate, high glucose (D5796-500ML, Sigma) supplemented with 10% donor bovine serum (16030074, Gibco), 1% L-Glutamine (25030-024, Gibco) and 1% Penicilin/streptomycin (15140-122, Gibco). Culture medium was changed after the 10^th^ day *in vitro* and twice per week afterwards. On reaching confluence, tanycytes were trypsinized and plated in plastic culture plate for internalization and secretion assays, on poly-L-lysine coated glass coverslips for secretion assays or on poly-L-lysine Ibidi glass bottom μ-Slide 8 well (80827, Ibidi) for live imaging.

### Primary tanycyte 2N4R Tau and Tau-565 internalization assay

Primary tanycytes were treated with either recombinant 2N4R human Tau (25 nM, 30 minutes) or Tau-565 (25 nM, 15 to 30 minutes) and Tau internalization was respectively analyzed using western blot and immunocytochemistry.

### Immunocytochemistry

For immunocytochemistry, coverslip seeded primary tanycytes, untreated and Tau-565 treated (25nM, 30 minutes), were washed three time with PBS and fixed in 4% PFA for 15 minutes at 4°C. After fixation, cells were washed with PBS and stored at 4° in PBS 0,1% azide. Coverslips of both untreated and Tau-565 tanycytes were incubated with primary antibodies (see Antibody Table) overnight in ICS at 4°C. The coverslips were washed three times in PBS prior to secondary antibody incubation in ICS for 1 hour at room temperature. Excess of secondary antibody was washed three times with PBS and cells were counterstained using DAPI before mounting in Mowiol.

### Western blot of primary tanycyte extracts

For western blot, confluent 6 well plates were treated with recombinant 2N4R human Tau (25nM, 30 minutes). After treatment, cells were washed five times with PBS and immediately frozen in dry-ice. Proteins extracts were prepared by scraping the cells in 1x RIPA buffer (20-188, Millipore) and homogenization by sonication. For each lane, 10μg of proteins in Laemlli buffer (1610747, Bio-Rad) was loaded in a 10% acrylamide gel. Migration was performed for 1h at 120V and followed by protein transfer onto a 0,45 μm nitrocellulose membrane for 1h at 100V. Immunolabeling of the membrane was performed using Tau5 mouse anti-Tau primary antibody (1:2,000, 606-320, ThermoFisher Scientific) or rabbit anti-GAPDH (1:5,000, G9545, Millipore) overnight in 5% milk TBS-Tween 0,05% at 4°C. Primary antibody was then washed three times with TBS-Tween 0,05% and incubated with HRP-coupled rabbit anti-mouse (P026002-2, Agilent) or HRP-coupled goat anti-rabbit (P044801-2, Agilent) secondary antibody for 1 hour at room temperature. Secondary antibodies in excess were washed three times with TBS-Tween 0,05% and membranes were incubated in SuperSignal™ West Dura Extended Duration Substrate (34076, Thermo Scientific). Chemiluminescent signal was detected using an Amersham ImageQuant 800 system (Cytiva).

### Primary tanycyte Tau-565 secretion assay

For tanycyte Tau-565 secretion assay, tanycytes were treated with Tau-565 (25nM, 30 minutes), washed five times with hot PBS and incubated with fresh Tau-565 free culture medium for 5, 15, 30 and 60 minutes. After the different secretion time points, the medium was recovered and immediately frozen in dry-ice. Tau concentration in the medium was measured by ELISA as described above.

### Live imaging of intracellular Tau transport

Prior to live imaging, primary tanycytes were incubated PKH67 green fluorescent cell linker (MIDI67-1KT, Sigma Aldrich) for 2 minutes to label cell membranes and then washed five times with culture medium. Then, they were treated with Tau-565 at 2μg.mL^−1^ (≈ 50nM) for 5 minutes and washed five times before live imaging acquisitions.

### Fluorescence microscopy

Microphotograph acquisitions were performed on a Zeiss AxioObserver Z1, associated with a spinning disk head (Yokogawa CSU-X1) and a camera (sCMOS Photometrics Prime 95B), confocal microscope using PLAN-APOCHROMAT 10x/0.45, 20x/0.8 and 63x/1.4, objectives under Zen 2.3 (Zeiss) software control. Live imaging acquisition were perfomed on the same microscope set up at a rate of 1,5 frames per second.

### Statistics

Results are given as mean ± standard deviation (s.d.). Data were excluded after ROUT method for outliers identification (Q=0,1%) for human data or when an objective experimental failure was observed for cell and animal experimentation. Studies were not randomized and investigators were not blind to treatment group. To test whether data followed a Gaussian distribution, a normality test was performed (Kolgomorov-Smirnov and Shapiro-Wilk tests). For normal distributions, two-sided unpaired *t*-test were used to compare two population and for multiple comparison one-way ANOVA followed by Tukey’s post hoc multiple-comparison test was used. For non-Gaussian distributions, Mann-Whitney test was used to compare to set of data and Kruskal-Wallis followed by Dunn’s post hoc test was used for multiple comparisons. Data analysis was performed using GraphPad Prism Software v8.1.1 (GraphPad). The threshold for significance was *P* < 0.05.

## Data Availability

All data produced in the present study are available upon reasonable request to the authors
Data collection and sharing for this project was funded by the Alzheimer Disease Neuroimaging Initiative (ADNI) (National Institutes of Health Grant U01 AG024904) and DOD ADNI (Department of Defense award number W81XWH-12-2-0012). ADNI is funded by the National Institute on Aging, the National Institute of Biomedical Imaging and Bioengineering, and through generous contributions from the following: AbbVie, Alzheimer Association; Alzheimer Drug Discovery Foundation; Araclon Biotech; BioClinica, Inc.; Biogen; Bristol-Myers Squibb Company; CereSpir, Inc.; Cogstate; Eisai Inc.; Elan Pharmaceuticals, Inc.; Eli Lilly and Company; EuroImmun; F. Hoffmann-La Roche Ltd and its affiliated company Genentech, Inc.; Fujirebio; GE Healthcare; IXICO Ltd.; Janssen Alzheimer Immunotherapy Research & Development, LLC.; Johnson & Johnson Pharmaceutical Research & Development LLC.; Lumosity; Lundbeck; Merck & Co., Inc.; Meso Scale Diagnostics, LLC.; NeuroRx Research; Neurotrack Technologies; Novartis Pharmaceuticals Corporation; Pfizer Inc.; Piramal Imaging; Servier; Takeda Pharmaceutical Company; and Transition Therapeutics. The Canadian Institutes of Health Research is providing funds to support ADNI clinical sites in Canada. Private sector contributions are facilitated by the Foundation for the National Institutes of Health (www.fnih.org). The grantee organization is the Northern California Institute for Research and Education, and the study is coordinated by the Alzheimer Therapeutic Research Institute at the University of Southern California. ADNI data are disseminated by the Laboratory for Neuro Imaging at the University of Southern California.

## ACKNOWLEDGMENT

Data collection and sharing for this project was funded by the Alzheimer’s Disease Neuroimaging Initiative (ADNI) (National Institutes of Health Grant U01 AG024904) and DOD ADNI (Department of Defense award number W81XWH-12-2-0012). ADNI is funded by the National Institute on Aging, the National Institute of Biomedical Imaging and Bioengineering, and through generous contributions from the following: AbbVie, Alzheimer’s Association; Alzheimer’s Drug Discovery Foundation; Araclon Biotech; BioClinica, Inc.; Biogen; Bristol-Myers Squibb Company; CereSpir, Inc.; Cogstate; Eisai Inc.; Elan Pharmaceuticals, Inc.; Eli Lilly and Company; EuroImmun; F. Hoffmann-La Roche Ltd and its affiliated company Genentech, Inc.; Fujirebio; GE Healthcare; IXICO Ltd.; Janssen Alzheimer Immunotherapy Research & Development, LLC.; Johnson & Johnson Pharmaceutical Research & Development LLC.; Lumosity; Lundbeck; Merck & Co., Inc.; Meso Scale Diagnostics, LLC.; NeuroRx Research; Neurotrack Technologies; Novartis Pharmaceuticals Corporation; Pfizer Inc.; Piramal Imaging; Servier; Takeda Pharmaceutical Company; and Transition Therapeutics. The Canadian Institutes of Health Research is providing funds to support ADNI clinical sites in Canada. Private sector contributions are facilitated by the Foundation for the National Institutes of Health (www.fnih.org). The grantee organization is the Northern California Institute for Research and Education, and the study is coordinated by the Alzheimer’s Therapeutic Research Institute at the University of Southern California. ADNI data are disseminated by the Laboratory for Neuro Imaging at the University of Southern California.

The authors are grateful to Meryem Tardivel and Antonino Bongiovanni (Confocal microscopy) from the BioImaging Center of Lille (BiCeL), and Julien Devassine (animal core facility) of the UMS2014-US41 for their expert technical support.

## AUTHOR CONTRIBUTION

F.S. and V.P. designed the study, analyzed data, prepared the figures, and wrote the manuscript. F.S. was involved in all aspect of study design, interpretations of the results and manuscript preparation. G.T. performed the light-sheet microscopy experiments. E. D. and C.D. participated in Tau-565 production and characterization. T.L. and F.P. monitored patients. J.D. and C-A.M. autopsied the patients and collected clinical samples. I.L and L.B. contributed material. T.L., S.R. P.C., P.G., Y-B.K., F.P., L.B., I.L., R.N. and M.S. were involved in the study design, interpretation of results, and preparation of the manuscript. S.R. edited the manuscript.

---

**Video 1: Tanycytes transport Tau-containing vesicles *in vitro***

Live imaging (1,5 frames per second, 1 minute of acquisition, 3x speed) of cultured primary tanycytes incubated with Tau-565 (red) and with plasma membranes stained in green.

**Video 2: Tanycytes throughout the median eminence take up Tau *in vivo***

3D light-sheet acquisition of the third ventricle of Tau-565 (white)-injected WT mice.

**Supplementary Table 1:**
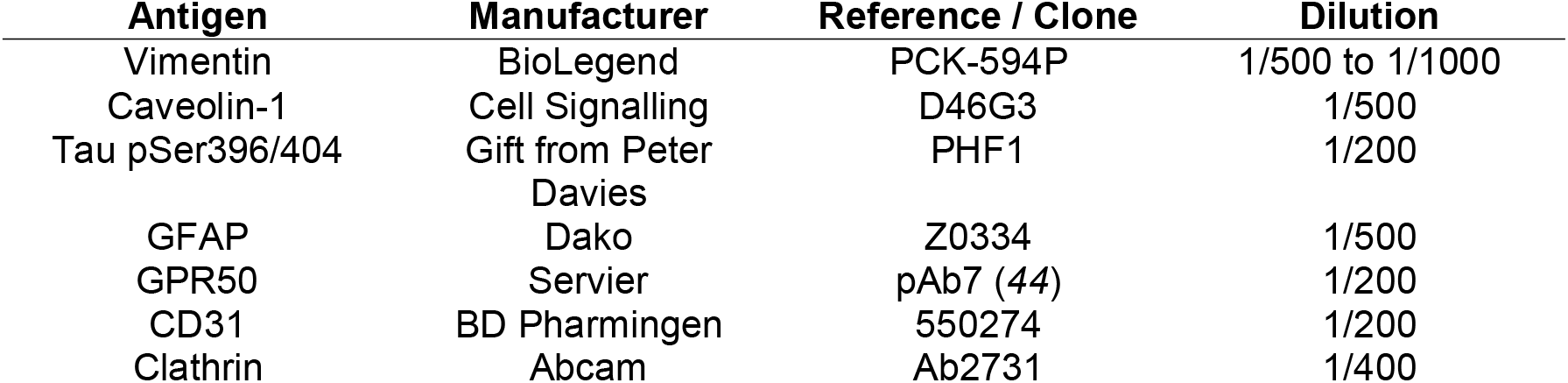
Antibodies for immunohistology on human and mice tissues.

| Antigen | Manufacturer | Reference / Clone | Dilution |
| --- | --- | --- | --- |
| Vimentin | BioLegend | PCK-594P | 1/500 to 1/1000 |
| Caveolin-1 | Cell Signalling | D46G3 | 1/500 |
| Tau pSer396/404 | Gift from Peter Davies | PHF1 | 1/200 |
| GFAP | Dako | Z0334 | 1/500 |
| GPR50 | Servier | pAb7 (44) | 1/200 |
| CD31 | BD Pharmingen | 550274 | 1/200 |
| Clathrin | Abcam | Ab2731 | 1/400 |

**Supplementary Figure 1:**
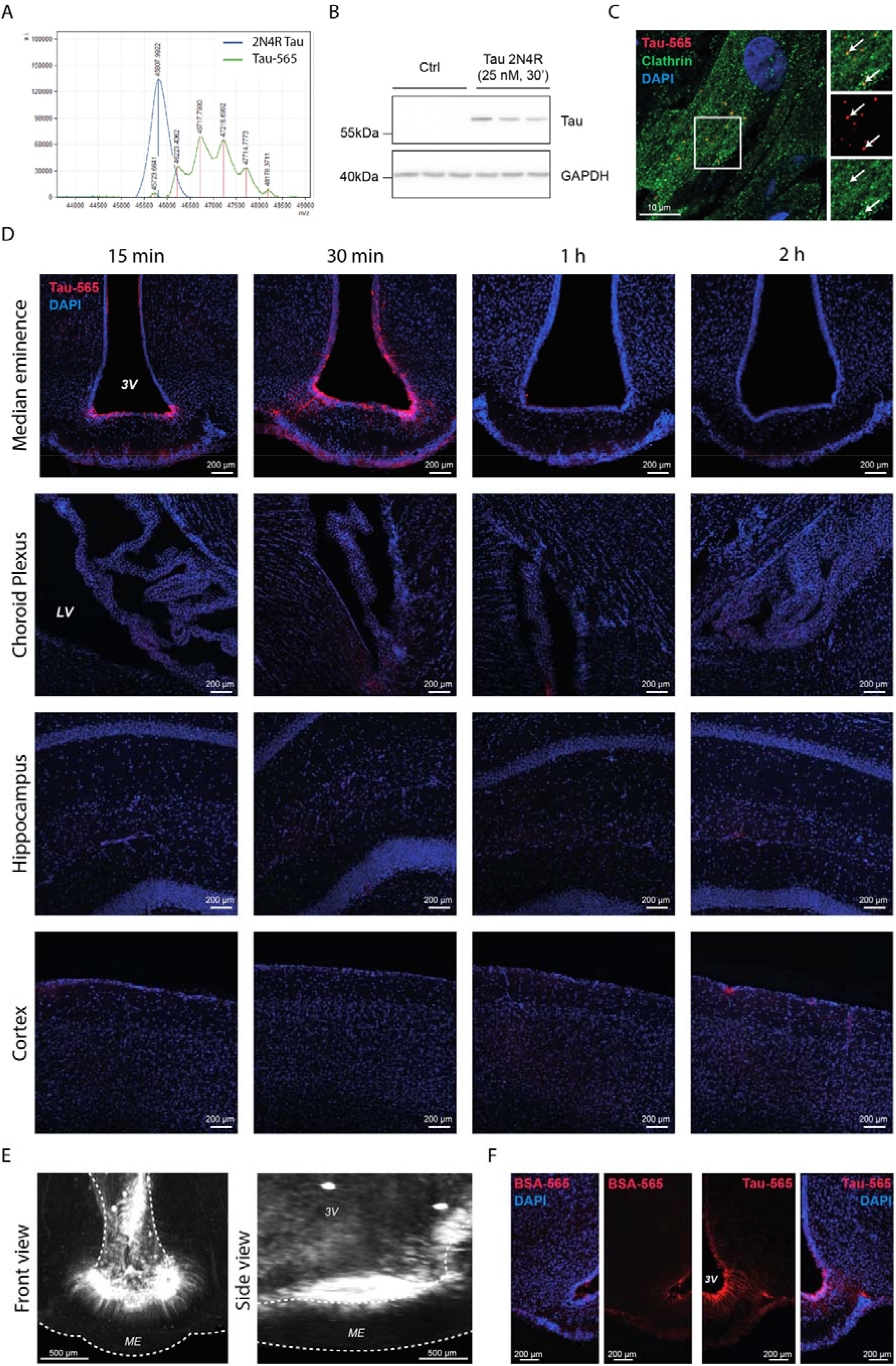
Tanycytes take up and secrete Tau *in vitro* and *in vivo*. A. Western blot for Tau protein in protein extracts of primary tanycyte cultures, untreated and treated with recombinant human 2N4R Tau, using the Tau5 antibody. B. Mass spectrometry plot of unconjugated recombinant 2N4R Tau and Atto-565-conjugated recombinant 2N4R Tau (Tau-565). Tau-565 peaks (noted by a red line) represent different Atto-565 conjugated 2N4R Tau species having up to 5 fluorophores per protein. C. Photomicrograph of cultured rat primary tanycytes treated or not with Tau-565 (red) and immunolabeled for clathrin (green). White arrows indicate double positive vesicles. D. Photomicrographs of the median eminence, choroid plexus, hippocampus and cortex of WT mice at different time points after Tau-565 (red) injection into the lateral ventricle. E. Coronal and sagittal reprojection of the third ventricle of Tau-565 (white)-injected WT mice: 3D light-sheet acquisition. F. Photomicrograph of the third ventricle and median eminence of WT mice injected with BSA-565 (red) (left panels) or Tau-565 (red) (right panels).

**Supplementary Figure 2:**
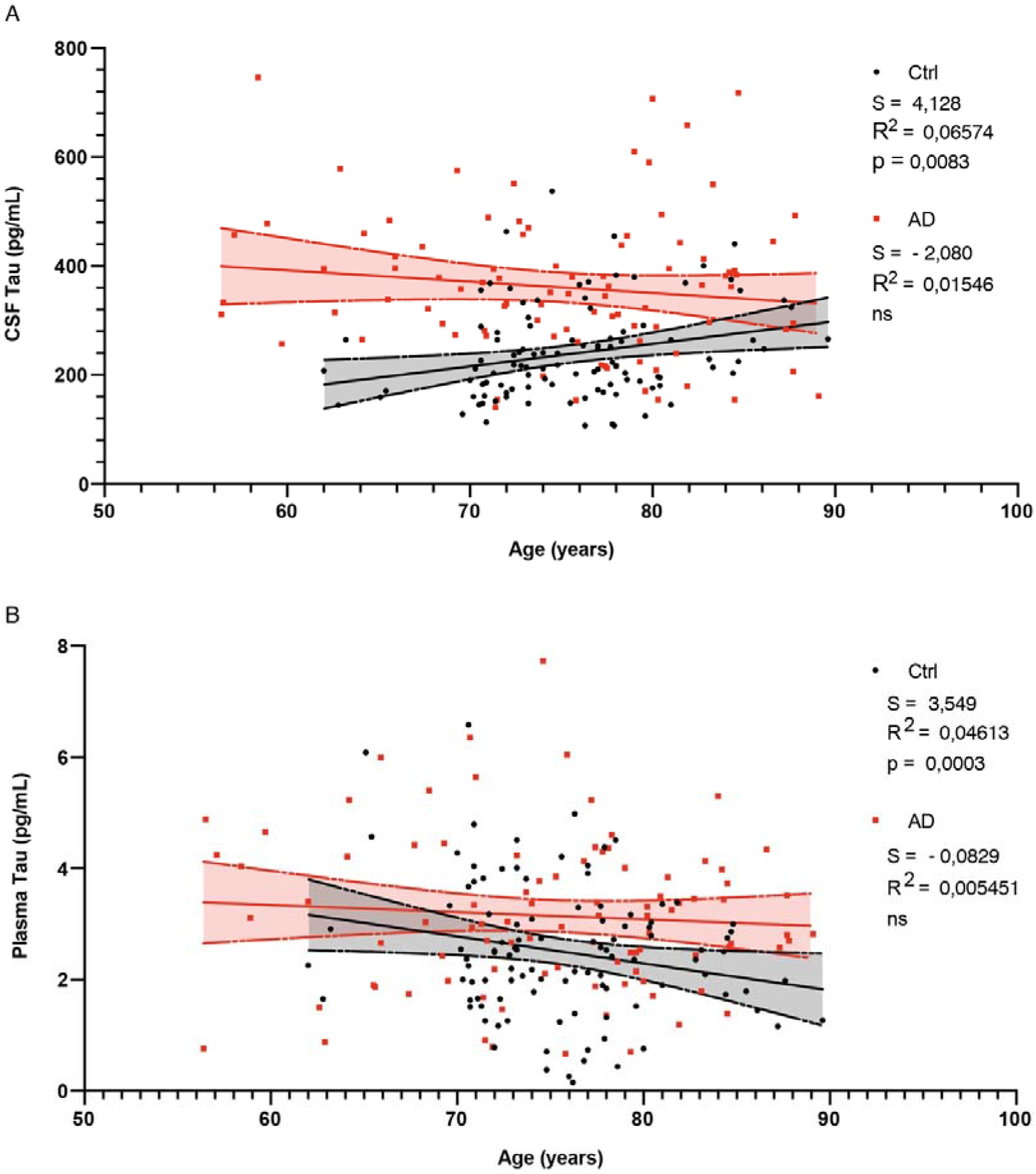
CSF and Plasma Tau changes with age in control and AD patients. A. CSF Tau concentration distribution in function of age in control (black, n=96) and AD (red, n=88) patients. Linear regression model and 95% confidence interval for control (black, dashed-lines and black shadow) and AD patients (red dashed-lines and red shadow). B. Plasma Tau concentration distribution in function of age in control (black, n=96) and AD (red, n=88) patients. Linear regression model and 95% confidence interval for control (black dashed-lines and black shadow) and AD patients (red dashed-lines and red shadow).

**Supplementary Figure 3:**
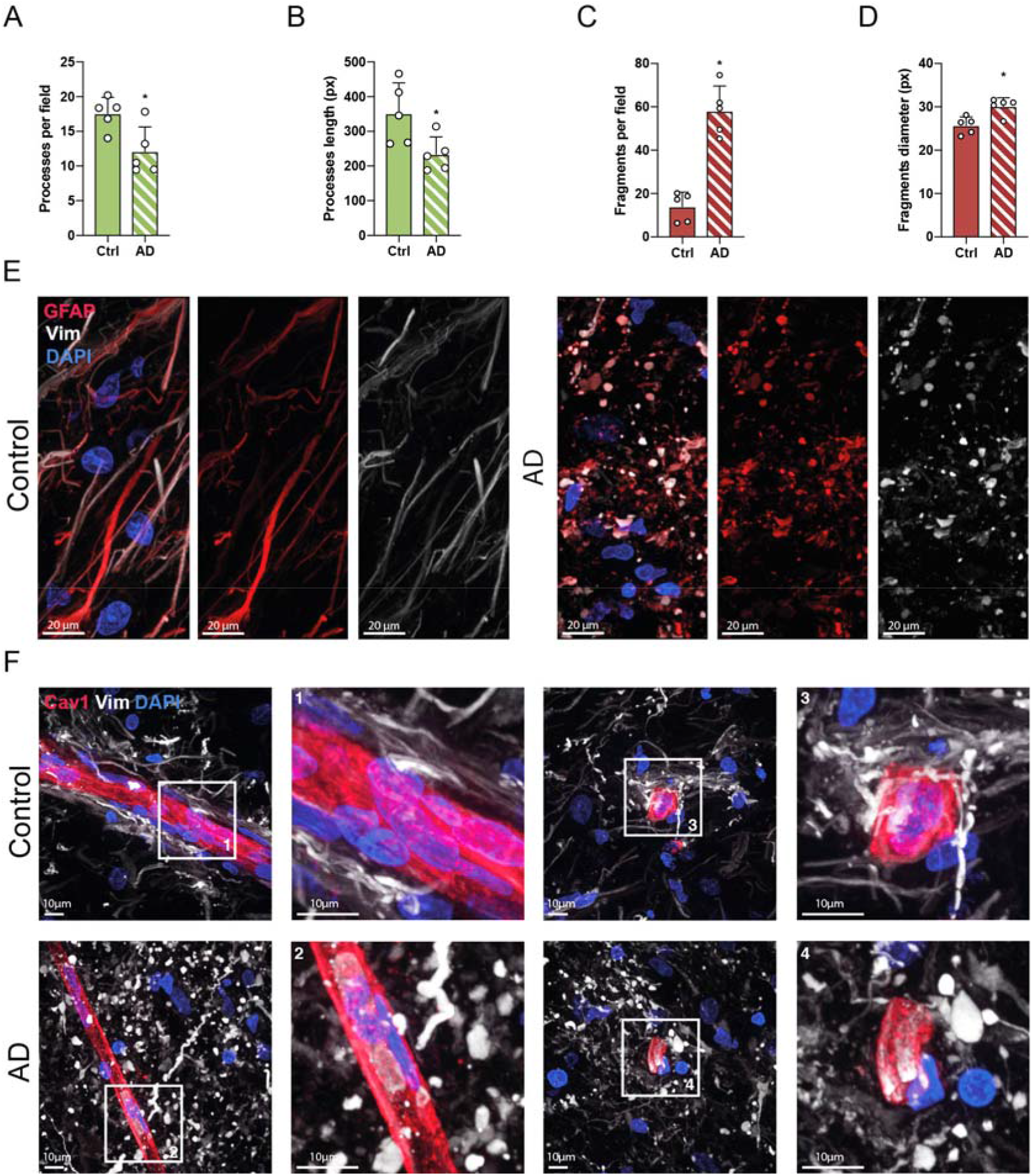
Tanycytes in AD patients are degraded and disorganized. A. Graph of tanycytic processes per field in control (n=5) and AD patients (n=5). Two-tailed Mann-Whitney U test (*U*=1, p=0.0317). Data are presented as means ± S.D. *, p<0.05. B. Graph of tanycytic process length in control (n=5) and AD patients (n=5). Two-tailed Mann-Whitney test (*U*=0, p=0.03). Data are presented as means ± S.D. *, p<0.05. C. Graph of tanycytic fragments per field in control (n=5) and AD patients (n=5). Two-tailed Mann-Whitney U test (*U*=0, p=0.0079). Data are presented as means ± S.D. *, p<0.05. D. Graph of tanycytic fragment diameter in control (n=5) and AD patients (n=5). Two-tailed Mann-Whitney U test (*U*=1, p=0.0159). Data are presented as means ± S.D. *, p<0.05. E. Photomicrographs of tanycyte cytoskeleton immunolabeling (vimentin in white, GFAP in red) in control (left panels) and AD patients (right panels). F. Photomicrograph of tanycyte-to-capillary contact in the infundibulum of control and AD patients immunolabeled for vimentin (white) to identify tanycytes and caveolin 1 (red) to identify capillaries. Capillaries parallel to the plane of the section are presented in the left panels and capillaries perpendicular to the plane of the section are presented in the right panels. Insets: higher magnification views of tanycyte-to-capillary contacts in control and AD patients.

## BIBLIOGRAPHY

1. 2021 Alzheimer’s disease facts and figures. Alzheimer’s & Dementia. n/a, doi:https://doi.org/10.1002/alz.12328.

2. G. G. Glenner, C. W. Wong, Alzheimer’s disease: Initial report of the purification and characterization of a novel cerebrovascular amyloid protein. Biochemical and Biophysical Research Communications. 120, 885–890 (1984).

3. J. P. Brion, A. M. Couck, E. Passareiro, J. Flament-Durand, Neurofibrillary tangles of Alzheimer’s disease: an immunohistochemical study. J Submicrosc Cytol. 17, 89–96 (1985).

4. I. Grundke-Iqbal, K. Iqbal, Y. C. Tung, M. Quinlan, H. M. Wisniewski, L. I. Binder, Abnormal phosphorylation of the microtubule-associated protein tau (tau) in Alzheimer cytoskeletal pathology. Proceedings of the National Academy of Sciences. 83, 4913–4917 (1986).

5. K. S. Kosik, C. L. Joachim, D. J. Selkoe, Microtubule-associated protein tau (tau) is a major antigenic component of paired helical filaments in Alzheimer disease. Proceedings of the National Academy of Sciences. 83, 4044–4048 (1986).

6. G. S. Bloom, Amyloid-β and Tau: The Trigger and Bullet in Alzheimer Disease Pathogenesis. JAMA Neurol. 71, 505 (2014).

7. J. P. Brion, H. Passareiro, J. Nunez, J. Flament-Durand, Mise en évidence immunologique de la protéine tau au niveau des lésions de dégénérescence neurofibrillaire de la maladie d’Alzheimer. Archives de biologie. 95, 229–235 (1985).

8. A. Delacourte, A. Defossez, Alzheimer’s disease: Tau proteins, the promoting factors of microtubule assembly, are major components of paired helical filaments. J Neurol Sci. 76, 173–186 (1986).

9. B. B. Holmes, J. L. Furman, T. E. Mahan, T. R. Yamasaki, H. Mirbaha, W. C. Eades, L. Belaygorod, N. J. Cairns, D. M. Holtzman, M. I. Diamond, Proteopathic tau seeding predicts tauopathy in vivo. Proc Natl Acad Sci U S A. 111, E4376–4385 (2014).

10. G. Chen, T. Xu, Y. Yan, Y. Zhou, Y. Jiang, K. Melcher, H. E. Xu, Amyloid beta: structure, biology and structure-based therapeutic development. Acta Pharmacol Sin. 38, 1205–1235 (2017).

11. J. M. Tarasoff-Conway, R. O. Carare, R. S. Osorio, L. Glodzik, T. Butler, E. Fieremans, L. Axel, H. Rusinek, C. Nicholson, B. V. Zlokovic, B. Frangione, K. Blennow, J. Ménard, H. Zetterberg, T. Wisniewski, M. J. de Leon, Clearance systems in the brain— implications for Alzheimer disease. Nat Rev Neurol. 11, 457–470 (2015).

12. I. F. Harrison, O. Ismail, A. Machhada, N. Colgan, Y. Ohene, P. Nahavandi, Z. Ahmed, Fisher, S. Meftah, T. K. Murray, O. P. Ottersen, E. A. Nagelhus, M. J. O’Neill, J. A. Wells, M. F. Lythgoe, Impaired glymphatic function and clearance of tau in an Alzheimer’s disease model. Brain. 143, 2576 (2020).

13. J. J. Iliff, M. J. Chen, B. A. Plog, D. M. Zeppenfeld, M. Soltero, L. Yang, I. Singh, R. Deane, M. Nedergaard, Impairment of Glymphatic Pathway Function Promotes Tau Pathology after Traumatic Brain Injury. The Journal of Neuroscience. 34, 16180 (2014).

14. K. Blennow, A. Wallin, H. Agren, C. Spenger, J. Siegfried, E. Vanmechelen, Tau protein in cerebrospinal fluid: a biochemical marker for axonal degeneration in Alzheimer disease? Mol Chem Neuropathol. 26, 231–245 (1995).

15. N. Andreasen, L. Minthon, E. Vanmechelen, H. Vanderstichele, P. Davidsson, B. Winblad, K. Blennow, Cerebrospinal fluid tau and Abeta42 as predictors of development of Alzheimer’s disease in patients with mild cognitive impairment. Neurosci Lett. 273, 5–8 (1999).

16. B. Olsson, R. Lautner, U. Andreasson, A. Öhrfelt, E. Portelius, M. Bjerke, M. Hölttä, C. Rosén, C. Olsson, G. Strobel, E. Wu, K. Dakin, M. Petzold, K. Blennow, H. Zetterberg, CSF and blood biomarkers for the diagnosis of Alzheimer’s disease: a systematic review and meta-analysis. The Lancet Neurology. 15, 673–684 (2016).

17. W. A. Banks, A. Kovac, P. Majerova, K. M. Bullock, M. Shi, J. Zhang, Tau Proteins Cross the Blood-Brain Barrier. Journal of Alzheimer’s Disease. 55, 411–419 (2017).

18. J. Wang, W.-S. Jin, X.-L. Bu, F. Zeng, Z.-L. Huang, W.-W. Li, L.-L. Shen, Z.-Q. Zhuang, Y. Fang, B.-L. Sun, J. Zhu, X.-Q. Yao, G.-H. Zeng, Z.-F. Dong, J.-T. Yu, Z. Hu, W. Song, H.-D. Zhou, J.-X. Jiang, Y.-H. Liu, Y.-J. Wang, Physiological clearance of tau in the periphery and its therapeutic potential for tauopathies. Acta Neuropathol. 136, 525–536 (2018).

19. N. Mattsson, H. Zetterberg, S. Janelidze, P. S. Insel, U. Andreasson, E. Stomrud, S. Palmqvist, D. Baker, C. A. T. Hehir, A. Jeromin, D. Hanlon, L. Song, L. M. Shaw, J. Q. Trojanowski, M. W. Weiner, O. Hansson, K. Blennow, Plasma tau in Alzheimer disease. Neurology. 87, 1827 (2016).

20. B. Engelhardt, L. Sorokin, The blood–brain and the blood–cerebrospinal fluid barriers: function and dysfunction. Semin Immunopathol. 31, 497–511 (2009).

21. P. Ciofi, M. Garret, O. Lapirot, P. Lafon, A. Loyens, V. Prévot, J. E. Levine, Brain-Endocrine Interactions: A Microvascular Route in the Mediobasal Hypothalamus. Endocrinology. 150, 5509–5519 (2009).

22. P. Ciofi, The arcuate nucleus as a circumventricular organ in the mouse. Neuroscience Letters. 487, 187–190 (2011).

23. H. M. Duvernoy, P.-Y. Risold, The circumventricular organs: an atlas of comparative anatomy and vascularization. Brain Res Rev. 56, 119–147 (2007).

24. V. Prevot, R. Nogueiras, M. Schwaninger, in Handbook of Clinical Neurology (Elsevier, 2021; https://linkinghub.elsevier.com/retrieve/pii/B9780128201077000161), vol. 180, pp. 253–273.

25. W. A. Banks, The blood-brain barrier as an endocrine tissue. Nat Rev Endocrinol. 15, 444–455 (2019).

26. A. Mullier, S. G. Bouret, V. Prevot, B. Dehouck, Differential distribution of tight junction proteins suggests a role for tanycytes in blood-hypothalamus barrier regulation in the adult mouse brain. J Comp Neurol. 518, 943–962 (2010).

27. V. Prevot, B. Dehouck, A. Sharif, P. Ciofi, P. Giacobini, J. Clasadonte, The Versatile Tanycyte: A Hypothalamic Integrator of Reproduction and Energy Metabolism. Endocrine Reviews. 39, 333–368 (2018).

28. M. Duquenne, C. Folgueira, C. Bourouh, M. Millet, A. Silva, J. Clasadonte, M. Imbernon, D. Fernandois, I. Martinez-Corral, S. Kusumakshi, E. Caron, S. Rasika, E. Deliglia, N. Jouy, A. Oishi, M. Mazzone, E. Trinquet, J. Tavernier, Y.-B. Kim, S. Ory, R. Jockers, M. Schwaninger, U. Boehm, R. Nogueiras, J.-S. Annicotte, S. Gasman, J. Dam, V. Prévot, Leptin brain entry via a tanycytic LepR–EGFR shuttle controls lipid metabolism and pancreas function. Nat Metab. 3, 1071–1090 (2021).

29. S. Gabery, C. G. Salinas, S. J. Paulsen, J. Ahnfelt-Rønne, T. Alanentalo, A. F. Baquero, S. T. Buckley, E. Farkas, C. Fekete, K. S. Frederiksen, W. F. J. Hogendorf, H. C. C. Helms, J. F. Jeppesen, L. M. John, C. Pyke, J. Nøhr, T. T. Lu, J. Polex-Wolf, V. Prevot, K. Raun, L. Simonsen, G. Sun, A. Szilvásy-Szabó, H. Willenbrock, A. Secher, L. B. Knudsen, Semaglutide lowers body weight in rodents via distributed neural pathways. JCI Insight. 5 (2020), doi:10.1172/jci.insight.133429.

30. G. Collden, E. Balland, J. Parkash, E. Caron, F. Langlet, V. Prevot, S. G. Bouret, Neonatal overnutrition causes early alterations in the central response to peripheral ghrelin. Mol Metab. 4, 15–24 (2015).

31. E. Balland, J. Dam, F. Langlet, E. Caron, S. Steculorum, A. Messina, S. Rasika, A. Falluel-Morel, Y. Anouar, B. Dehouck, E. Trinquet, R. Jockers, S. G. Bouret, V. Prévot, Hypothalamic tanycytes are an ERK-gated conduit for leptin into the brain. Cell Metab. 19, 293–301 (2014).

32. M. Porniece Kumar, A. L. Cremer, P. Klemm, L. Steuernagel, S. Sundaram, A. Jais, A. C. Hausen, J. Tao, A. Secher, T. Å. Pedersen, M. Schwaninger, F. T. Wunderlich, B. B. Lowell, H. Backes, J. C. Brüning, Insulin signalling in tanycytes gates hypothalamic insulin uptake and regulation of AgRP neuron activity. Nat Metab. 3, 1662–1679 (2021).

33. T. K. Patel, L. Habimana-Griffin, X. Gao, B. Xu, S. Achilefu, K. Alitalo, C. A. McKee, P. W. Sheehan, E. S. Musiek, C. Xiong, D. Coble, D. M. Holtzman, Dural lymphatics regulate clearance of extracellular tau from the CNS. Mol Neurodegener. 14, 11 (2019).

34. L. Veys, J. Van houcke, J. Aerts, S. Van Pottelberge, M. Mahieu, A. Coens, R. Melki, D. Moechars, L. De Muynck, L. De Groef, Absence of Uptake and Prion-Like Spreading of Alpha-Synuclein and Tau After Intravitreal Injection of Preformed Fibrils. Front Aging Neurosci. 12, 614587 (2021).

35. M. Slezak, A. Grosche, A. Niemiec, N. Tanimoto, T. Pannicke, T. A. Münch, B. Crocker, P. Isope, W. Härtig, S. C. Beck, G. Huber, G. Ferracci, M. Perraut, M. Reber, M. Miehe, V. Demais, C. Lévêque, D. Metzger, K. Szklarczyk, R. Przewlocki, M. W. Seeliger, D. Sage-Ciocca, J. Hirrlinger, A. Reichenbach, S. Reibel, F. W. Pfrieger, Relevance of Exocytotic Glutamate Release from Retinal Glia. Neuron. 74, 504–516 (2012).

36. G. G. Schiavo, F. Benfenati, B. Poulain, O. Rossetto, P. P. de Laureto, B. R. DasGupta, C. Montecucco, Tetanus and botulinum-B neurotoxins block neurotransmitter release by proteolytic cleavage of synaptobrevin. Nature. 359, 832–835 (1992).

37. M. Banerjee, S. Joshi, J. Zhang, C. L. Moncman, S. Yadav, B. A. Bouchard, B. Storrie, S. W. Whiteheart, Cellubrevin/vesicle-associated membrane protein-3–mediated endocytosis and trafficking regulate platelet functions. Blood. 130, 2872–2883 (2017).

38. J. F. Caro, J. W. Kolaczynski, M. R. Nyce, J. P. Ohannesian, I. Opentanova, W. H. Goldman, R. B. Lynn, P.-L. Zhang, M. K. Sinha, R. V. Considine, Decreased cerebrospinal-fluid/serum leptin ratio in obesity: a possible mechanism for leptin resistance. The Lancet. 348, 159–161 (1996).

39. T. Skillbäck, L. Delsing, J. Synnergren, N. Mattsson, S. Janelidze, K. Nägga, L. Kilander, R. Hicks, A. Wimo, B. Winblad, O. Hansson, K. Blennow, M. Eriksdotter, H. Zetterberg, CSF/serum albumin ratio in dementias: a cross-sectional study on 1861 patients. Neurobiology of Aging. 59, 1–9 (2017).

40. N. R. Barthélemy, K. Horie, C. Sato, R. J. Bateman, Blood plasma phosphorylated-tau isoforms track CNS change in Alzheimer’s disease. Journal of Experimental Medicine. 217, e20200861 (2020).

41. Y. Guo, Y.-Y. Huang, X.-N. Shen, S.-D. Chen, H. Hu, Z.-T. Wang, L. Tan, J.-T. Yu, the Alzheimer’s Disease Neuroimaging Initiative, Characterization of Alzheimer’s tau biomarker discordance using plasma, CSF, and PET. Alzheimer’s Research & Therapy. 13, 93 (2021).

42. M. W. Weiner, D. P. Veitch, P. S. Aisen, L. A. Beckett, N. J. Cairns, R. C. Green, D. Harvey, C. R. Jack, W. Jagust, E. Liu, J. C. Morris, R. C. Petersen, A. J. Saykin, M. E. Schmidt, L. Shaw, J. A. Siuciak, H. Soares, A. W. Toga, J. Q. Trojanowski, Alzheimer’s Disease Neuroimaging Initiative, The Alzheimer’s Disease Neuroimaging Initiative: A review of papers published since its inception. Alzheimer’s &amp; Dementia. 8 (2012), doi:10.1016/j.jalz.2011.09.172.

43. S. Berg, D. Kutra, T. Kroeger, C. N. Straehle, B. X. Kausler, C. Haubold, M. Schiegg, J. Ales, T. Beier, M. Rudy, K. Eren, J. I. Cervantes, B. Xu, F. Beuttenmueller, A. Wolny, C. Zhang, U. Koethe, F. A. Hamprecht, A. Kreshuk, ilastik: interactive machine learning for (bio)image analysis. Nat Methods. 16, 1226–1232 (2019).

44. A. Sidibe, A. Mullier, P. Chen, M. Baroncini, J. A. Boutin, P. Delagrange, V. Prevot, R. Jockers, Expression of the orphan GPR50 protein in rodent and human dorsomedial hypothalamus, tanycytes and median eminence. Journal of Pineal Research. 48, 263–269 (2010).

45. S.-S. Yoon, S. A. Jo, Mechanisms of Amyloid-β Peptide Clearance: Potential Therapeutic Targets for Alzheimer’s Disease. Biomolecules & Therapeutics. 20, 245 (2012).

46. K. A. Bates, G. Verdile, Q.-X. Li, D. Ames, P. Hudson, C. L. Masters, R. N. Martins, Clearance mechanisms of Alzheimer’s amyloid-β peptide: implications for therapeutic design and diagnostic tests. Mol Psychiatry. 14, 469–486 (2009).

47. A. S. Chesser, S. M. Pritchard, G. V. W. Johnson, Tau Clearance Mechanisms and Their Possible Role in the Pathogenesis of Alzheimer Disease. Frontiers in Neurology. 4 (2013), doi:10.3389/fneur.2013.00122.

48. I. F. Harrison, O. Ismail, A. Machhada, N. Colgan, Y. Ohene, P. Nahavandi, Z. Ahmed, Fisher, S. Meftah, T. K. Murray, O. P. Ottersen, E. A. Nagelhus, M. J. O’Neill, J. A. Wells, M. F. Lythgoe, Impaired glymphatic function and clearance of tau in an Alzheimer’s disease model. Brain. 143, 2576–2593 (2020).

49. S.-H. Xin, L. Tan, X. Cao, J.-T. Yu, L. Tan, Clearance of Amyloid Beta and Tau in Alzheimer’s Disease: from Mechanisms to Therapy. Neurotox Res. 34, 733–748 (2018).

50. D. M. Barten, G. W. Cadelina, N. Hoque, L. B. DeCarr, V. L. Guss, L. Yang, S. Sankaranarayanan, P. D. Wes, M. E. Flynn, J. E. Meredith, M. K. Ahlijanian, C. F. Albright, Tau transgenic mice as models for cerebrospinal fluid tau biomarkers. J Alzheimers Dis. 24 Suppl 2, 127–141 (2011).

51. M. Zoli, F. Ferraguti, A. Frasoldati, G. Biagini, L. F. Agnati, Age-related alterations in tanycytes of the mediobasal hypothalamus of the male rat. Neurobiology of Aging. 16, 77–83 (1995).

52. A. C. M. Koopman, M. Taziaux, J. Bakker, Age-related changes in the morphology of tanycytes in the human female infundibular nucleus/median eminence. J Neuroendocrinol. 29 (2017), doi:10.1111/jne.12467.

53. A. Chen, R. O. Akinyemi, Y. Hase, M. J. Firbank, M. N. Ndung’u, V. Foster, L. J. L. Craggs, K. Washida, Y. Okamoto, A. J. Thomas, T. M. Polvikoski, L. M. Allan, A. E. Oakley, J. T. O’Brien, K. Horsburgh, M. Ihara, R. N. Kalaria, Frontal white matter hyperintensities, clasmatodendrosis and gliovascular abnormalities in ageing and poststroke dementia. Brain. 139, 242–258 (2016).

54. H. Tomimoto, I. Akiguchi, H. Wakita, T. Suenaga, S. Nakamura, J. Kimura, Regressive changes of astroglia in white matter lesions in cerebrovascular disease and Alzheimer’s disease patients. Acta Neuropathol. 94, 146–152 (1997).

55. Y. Hase, K. Horsburgh, M. Ihara, R. N. Kalaria, White matter degeneration in vascular and other ageing-related dementias. J Neurochem. 144, 617–633 (2018).

56. N. Daschil, C. Humpel, Green-Fluorescent Protein+ Astrocytes Attach to Beta-Amyloid Plaques in an Alzheimer Mouse Model and Are Sensitive for Clasmatodendrosis. Frontiers in Aging Neuroscience. 8 (2016), doi:10.3389/fnagi.2016.00075.

57. D. Lana, F. Ugolini, M. G. Giovannini, Space-Dependent Glia–Neuron Interplay in the Hippocampus of Transgenic Models of β-Amyloid Deposition. International Journal of Molecular Sciences. 21 (2020), doi:10.3390/ijms21249441.

58. D. Lana, L. Iovino, D. Nosi, G. L. Wenk, M. G. Giovannini, The neuron-astrocyte-microglia triad involvement in neuroinflammaging mechanisms in the CA3 hippocampus of memory-impaired aged rats. Experimental Gerontology. 83, 71–88 (2016).

59. S. P. Raikwar, S. M. Bhagavan, S. B. Ramaswamy, R. Thangavel, I. Dubova, G. P. Selvakumar, M. E. Ahmed, D. Kempuraj, S. Zaheer, S. Iyer, A. Zaheer, Are Tanycytes the Missing Link Between Type 2 Diabetes and Alzheimer’s Disease? Mol Neurobiol. 56, 833–843 (2019).

60. M. Ishii, C. Iadecola, Metabolic and Non-Cognitive Manifestations of Alzheimer’s Disease: The Hypothalamus as Both Culprit and Target of Pathology. Cell Metab. 22, 761–776 (2015).

61. C. Danis, C. Despres, L. M. Bessa, I. Malki, H. Merzougui, I. Huvent, H. Qi, G. Lippens, F.-X. Cantrelle, R. Schneider, X. Hanoulle, C. Smet-Nocca, I. Landrieu, Nuclear Magnetic Resonance Spectroscopy for the Identification of Multiple Phosphorylations of Intrinsically Disordered Proteins. Journal of Visualized Experiments □: JoVE (2016), doi:10.3791/55001.

62. H. Müller-Fielitz, M. Stahr, M. Bernau, M. Richter, S. Abele, V. Krajka, A. Benzin, J. Wenzel, K. Kalies, J. Mittag, H. Heuer, S. Offermanns, M. Schwaninger, Tanycytes control the hormonal output of the hypothalamic-pituitary-thyroid axis. Nat Commun. 8, 484 (2017).

63. M. Belle, D. Godefroy, G. Couly, S. A. Malone, F. Collier, P. Giacobini, A. Chédotal, Tridimensional Visualization and Analysis of Early Human Development. Cell. 169, 161–173.e12 (2017).

64. V. Prevot, A. Cornea, A. Mungenast, G. Smiley, S. R. Ojeda, Activation of erbB-1 Signaling in Tanycytes of the Median Eminence Stimulates Transforming Growth Factor βRelease via Prostaglandin E2 Production and Induces Cell Plasticity. J neurosci. 19:10622-10632 (2003).

